# Study protocol for a Critical Realist pilot cluster-randomised controlled trial of a whole-school-based mindfulness intervention (SBMI) promoting child and adolescent mental wellbeing in Rwanda and Ethiopia

**DOI:** 10.1101/2023.05.10.23289769

**Authors:** Pamela Abbott, Lucia D’Ambruoso, Mahlet Yared, Paul McNamee, Tsion Hailu, Wenceslas Nzabalirwa

## Abstract

****Background**:** This research uses a critical realist approach to understand how and why school-based mindfulness interventions designed to promote child and adolescent mental wellbeing work or do not. Poor mental wellbeing is the leading cause of illness among children and adolescents in sub-Saharan Africa, with an estimated prevalence of 1 in 7. There is evidence that school-based mindfulness interventions promote child and adolescent wellbeing. Still, few interventions have been trialled in SSA, and none have examined how and why mindfulness interventions work.

**Methods:** Using a Critical Realist pilot cluster-randomised controlled trial; we will evaluate a school-based mindfulness intervention compared to the regular curriculum over one school year. Incorporating mindfulness into the primary school curriculum will provide proactive reach to all CA. The intervention will be codesigned by teacher educators, schoolteachers, policy actors and community members, including CAs. In each country, three schools will be selected and randomised as pilot, intervention, or control/waitlist. The mindfulness intervention will be integrated into the school curriculum and taught to all children attending the schools by classroom teachers trained to deliver it. Quantitative research will be used to measure the outcome of the intervention, and qualitative research to answer how and why questions. The primary outcome is improved mental wellbeing of pupils measured by the Acholi Psychological Assessment Instrument. Secondary outcomes will include subjective quality of life, school 'climate', school performance, and satisfaction with school. Pupils, teachers and main carers in intervention and control/waitlist schools will fill in questionnaires before and after the intervention and process evaluation will be carried out in intervention schools. The cost-effectiveness of the mindfulness intervention will be assessed.

**Discussion:** The evaluation will provide new interdisciplinary knowledge, and methods, to understand and sustainable impacts on CA mental wellbeing in these settings. Independent research and intervention teams will run the trial.

**Registration of Project**: Research Registry 8799, Mar 31 2023

## Introduction: Background and Rationale

This protocol sets out a programme of interdisciplinary, applied health research to investigate the potential for a school-based mindfulness intervention (SBMI) to promote the mental wellbeing of children and adolescents (CAs) in Rwanda and Ethiopia(1). It is designed as a pilot critical realist cluster control trial (CRcCT) of a culturally adapted SBMI. Such interventions are relatively inexpensive, non-stigmatising and inclusive and can be integrated into the school curriculum and taught by teachers after pre-or in-service training (2). A CRcCT enables the evaluation to go beyond measuring the relationship between the dependent and experimental variables and provides an understanding of how outcomes are influenced by deeper mechanisms, structures and processes, including underlying power dynamics, social norms, and cultural beliefs in a given context (3–7). The design enables us to enumerate the outcomes, develop and test hypotheses about the mechanisms embedded in the intervention and its context, and uncover how the CAs involved respond to the resources offered by the mindfulness intervention in the context (3). The evaluation will provide a more comprehensive and nuanced understanding of the intervention and enable us to identify ways to improve its effectiveness and sustainability. It will provide policy stakeholders with information about the intervention, what works and what conditions need to be in place for it to work. This is important for replicating the intervention and identifying possible generalisable causal pathways through replication in other contexts enabling the identification of demi-regularities, and semi-predictable patterns of programme functioning. Globally, mental health problems are the leading cause of illness in CAs (8), including LMICs (9). In Sub-Saharan Africa (SSA), for example, an estimated one in seven CAs have a mental health challenge (10), with even higher rates for adolescents (11). CAs in SSA face health and socioeconomic challenges that increase vulnerability to poor mental wellbeing (10,12,13). High rates of HIV, conflict and violence, fragile states, natural disasters, forced migration and discrimination are significant drivers of poor mental health (14). Poor mental wellbeing is associated with low educational attainment and increased involvement in risky behaviours, with mental health problems persisting into adulthood (15–17). However, the mental health needs of CAs are under-recognised. There is a shortage of evidence on promoting CAs’ mental wellbeing (10), a lack of policies and community awareness, poor mental health literacy, high levels of stigma and few interventions to reduce or detect CAs at risk (14,18). There is a widening treatment gap with few specialist services or qualified mental health workers and poor service delivery (12,13,19,20).

Rwanda and Ethiopia, where we will implement our programme, are aid-dependent, least-developed countries in East Africa. Child poverty is high; in Rwanda, 60% of children lived in absolute poverty (less than $1.95ppp a day) in 2017 and 26% in Ethiopia (21). Both countries are fragile, post-conflict states with CAs facing significant challenges, Ethiopia is ranked 153, and Rwanda ranked 130 out of 180 countries on the Child Flourishing Index (18). Primary school education is mandatory, and participation rates are high. However, children make slow progress through school, primary school completion rates are low, and most of those that do complete are over-age (22,23). Violence is part of the reality of everyday life for CAs, with physical punishment seen as normatively acceptable and used frequently (24–28). There is little reliable evidence on the extent of mental health problems among CAs. In Rwanda, the national mental health survey found a prevalence rate for mental disorders among those aged 14-18 of 10% (29). In Ethiopia, the CAs prevalence rate has been estimated to be between 12% and 25% (30). Both countries lack a clear plan for promoting CAs’ mental wellbeing, a lack of funding, an acute shortage of qualified mental health workers and no specialist services for CAs (31–37).

Mental wellbeing has an impact on CAs’ quality of life, their cognitive development, their attainment and physical health, as well as their mental wellbeing as adults (38,39). It is about more than the absence of mental illness; it is about their positive functioning. Mental wellbeing influences how CAs cope with stress, trauma, and physical ill-health and is influenced by their resilience, relationships, and the broader socioeconomic, cultural, and environmental conditions in which they live. However, it is possible to improve mental wellbeing through prevention and early interventions, including school-based ones (40–44). Such interventions focus on enhancing capabilities and preventing risky behaviour, are not stigmatising, and reach a high proportion of CAs (12,45–47). They are low-cost, reduce the treatment gap, and recognise the importance of working collaboratively with students, families, communities and teachers (48). Research has also shown that the cultural translation of interventions is essential and possible (49,50) with teachers trained to teach them (51). However, there is an acute need to strengthen the evidence base for low-and middle-income countries (LMICs), given the potential for low-cost whole-school interventions to have a high impact in settings of complex, multidimensional and intergenerational hardship, disadvantage and exclusion (52,53).

Mindfulness has been identified in HICs as an effective intervention for promoting CAs’ mental wellbeing, as evidenced by systematic reviews and meta-analyses (42,43,54), with interventions taking a school-based universal prevention approach being more likely to have a positive impact (38,54). Mindfulness positively affects several outcome domains, including improving cognitive performance, reducing stress, and increasing resilience and socio-emotional skills. There have been calls for the broad introduction of mindfulness in schools (43,55,56). Mindfulness interventions have also been shown to reduce teachers’ stress levels and improve their mental wellbeing and relationships with students making classroom environments more conducive to student learning (56–59). They can be integrated into the school timetable and delivered by teachers with pre-or in-service training.

Evidence on the efficacy of mindfulness in LMICs is less robust but suggests that it positively impacts CA mental wellbeing and decreases teachers’ stress when adapted to be culturally appropriate (49,60–64). There has been little research on SSA, and only one SBMI was evaluated using a quasi-experimental method in Northern Uganda (61). The evaluation found that adapting an intervention and training teachers was possible. It significantly decreased pupils’ depressive symptoms, reduced anger, hostility and feelings of rejection, and improved academic grades. Furthermore, a systematic review of research on task shifting in SSA concluded that teachers could be a valuable and cost-effective resource for mental health service delivery provided policies and training are in place, and teachers have been trained to deliver interventions (65).

The mindfulness intervention for this project (66) will be developed during Phase 1 of the project. In year 1, seven teacher educators, four from Rwanda and three from Ethiopia, will be educated in mindfulness by experienced mindfulness teacher educators from Scotland. In year 2, 20 teacher mindfulness champions, ten from Ethiopia and ten from Rwanda will be trained in mindfulness. They will work with the teacher educators to develop a culturally appropriate whole-school mindfulness intervention for schools in Rwanda and Ethiopia (67). Participatory Action Research with parents and CAs, interviews with policy stakeholders, and findings from literature reviews will inform the design of the intervention. The intervention will enable schools to create a culture of mindfulness that supports students’ wellbeing, academic success, and teachers’ personal and professional growth (56). The training will have three core interrelated elements: (1) intention, knowing what we are doing and why we are doing it; (2) attention, attending fully to the present moment; and (3) attitude, being open, kind and curious (68). Mindfulness practice will be integrated into the school day and curriculum, and CAs will be encouraged to share their experiences with parents and family members. The schools will promote mindful communication encouraging teachers and CAs to listen attentively, speak truthfully, use nonviolent communication techniques to resolve conflicts and foster a supportive, non-judgmental environment. They will also promote CAs to understand and appreciate the perspectives of others and to show kindness and care towards themselves and others.

To provide more evidence on the efficacy of SBMIs in LICs in SSA, the project will investigate the impact of a culturally-appropriate SBMI on the mental wellbeing of CAs in Ethiopia and Rwanda using a CR approach to determine what makes the programmes work, how, where, with whom and to what extent (69–71). The aim is to evaluate a culturally appropriate SBMI developed with stakeholders to promote CAs’ mental wellbeing.

The objectives of the project are:

(1) to enumerate the status of the mental wellbeing of CAs before and after the intervention;
(2) to identify the underlying context and intervention mechanisms explaining the effects of the mindfulness intervention and compare these with those identified in the literature;
(3) to examine how different CAs respond to the mindfulness intervention;
(4) to determine if the intervention is culturally acceptable and affordable?

## Methods and Analysis

### Introduction

This protocol sets out the design of a study which adopts a critical realist design. The protocol was guided by the Spirit 2013 checklist for Clinical Trial Protocols and CONSORT 2010 Checklist of information when reporting a pilot or feasibility trial, as recommended by Thabane and Lancaster (72). Critical realist trials follow the steps in the Medical Research Council’s framework for evaluating complex healthcare interventions (3).

The project will be implemented in two phases (Figure 1). Phase 1 will last 18 months, and the main activities of Phase 2 will last 30 months. The findings from Phase 1 will be used to inform the intervention design and refine the Programme Theory, which will be tested in Phase 2. Phase 2 will be an impact evaluation of the whole school mindfulness intervention. The intervention will be delivered in the intervention schools for nine months in year 3 (a school year) and in the control/waitlist schools in year 4. Before the mindfulness intervention is delivered in year three a pilot of the intervention and the research tools will be carried out in a feasibility school in each country.

**Figure 1:**
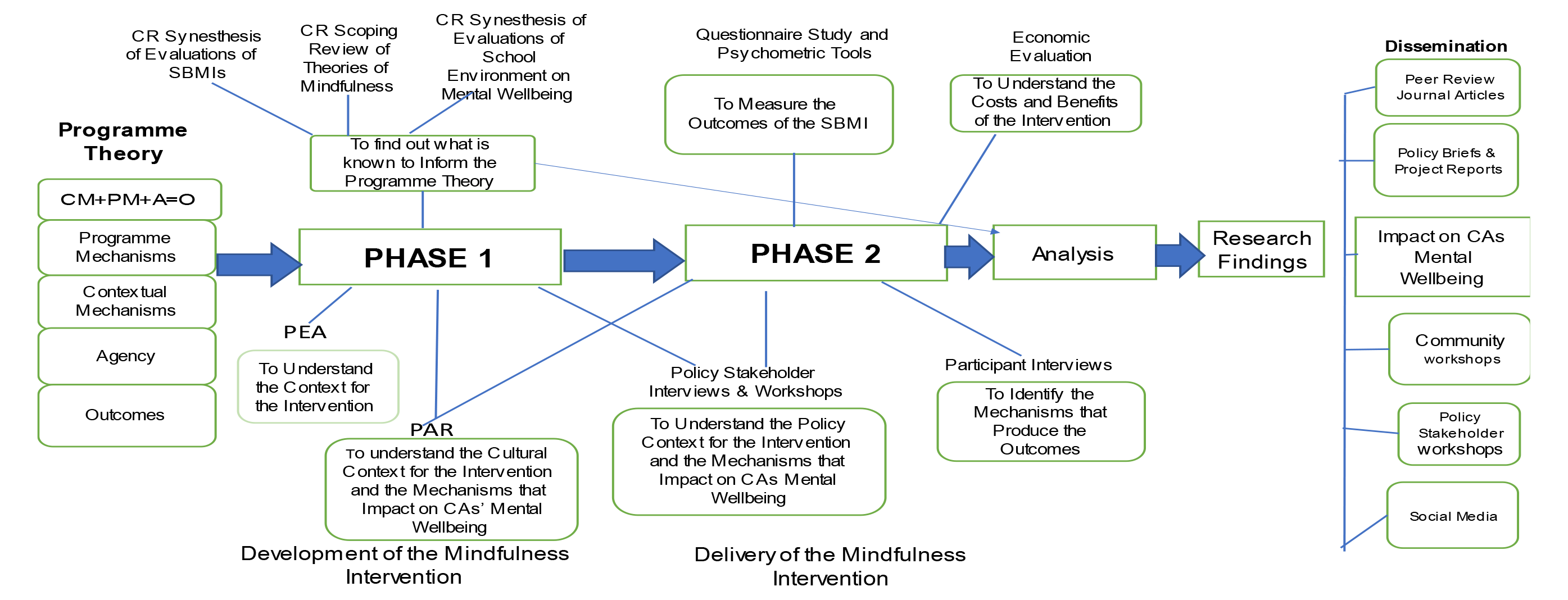
Study Illustration: Phase 1 and Phase 2 Methods and Outputs.

### Project Design

The overarching methodology is critical realism, a theory-driven form of case study evaluation that recognises that not every intervention will work for every person in the same way (73) and a rejection of the assumption that the effectiveness of an intervention is based only on its inherent qualities. The basic tenets of CR are ontological realism, epistemic relativism, judgemental rationality, a separation between structure and agency, and a commitment to social justice to understand how to improve people’s lives. Reality exists independently of our knowledge, with our understanding of it is constructed through social and cultural contexts. For CR, there are three overlapping ontological domains, the empirical, the actual and the real (74). The empirical consists of what reflexive agents observe, experience, and understand; the actual is where events generated by mechanisms and structures happen whether we experience them or not, and the real, which pre-exists us, is beyond the consciousness of human beings and is where the enduring mechanisms and structures exist that can produce events in the world. People are conditioned by the context in which they live, and this constrains their agency, but it is agency that brings about social change (75).

The project’s research design follows Bhaskar’s Critical Realist RRRIREI(C) (resolution, abductive redescription, retroduction, retrodiction, elimination, identification of antecedents and correction) model, which sets out the steps for social scientists to conduct applied explanatory research (Table 1) (76) combined with Margaret Archer’s Morphogenic approach (75). Bhaskar’s model provides a framework for interdisciplinary research, starting by determining which disciplines can contribute to the research. The aim is to move from disciplinary research and theorising to develop interdisciplinary theories (explanations) for the findings.

**Table 1:** Stages of Explanatory Critical Realist Applied Research.

| Planning and<br>Multi-disciplinary Phase |  | Cross-Phase<br>Phase |  |  | Interdisciplinary<br>Phase |  |
| --- | --- | --- | --- | --- | --- | --- |
| Resolution | Redescription | Retroduction | Retrodiction | Elimination | Identification | Correction |
| Identification of the levels that are included.<br>Biological/<br>Individual<br>Social-Interaction<br>Socioeconomic | Identification of disciplines and using abduction to develop theoretical explanations for the impact of the intervention on children's mental wellbeing | Identifying what properties are necessary for a phenomenon to exist | Identifying how different mechanisms reinforce, moderate, and condition one another to affect the outcome | Elimination of alternative theoretical explanations . | Identify the most comprehensive interdisciplinary explanation for how mindfulness promotes children's mental wellbeing | Identifying gaps or errors in existing theory in the light of the explanation developed by the programme. |
| Identifying disciplines that research at different levels and carrying out the research based on disciplinary expertise. | Develop disciplinary theories to explain the impact.<br>Reductionist/atomistic explanations. | Developing or revising existing hypotheses based on observation of their effects by identifying the underlying causal mechanisms that caused these effects | Develop trans-factual theories integrating the disciplinary explanations . | Use judgemental rationality to eliminate theories that lack explanatory power | Use judgemental rationality to identify the most comprehensive explanation | Iterative refinement of theory to account for new information to correct errors |

Based on Bhaskar, R. (2016). *Enlightened Common Sense: The Philosophy of Critical Realism*. Routledge: London and New York

Margaret Archer’s Morphogenic approach enables us to understand how an intervention works (Figure 2). She argues that structures shape human behaviour and social interaction while recognising that structures are not fixed or immutable but can be transformed through human agency. She argues that it is necessary to distinguish between agency and structure and between social structure and culture. People are conditioned by the context in which they are born but can change the social structure and culture through agency. The outcome of an intervention results from people’s interpretations and responses to the intervention in an open system where there is a complex interaction of a plurality of mechanisms, in addition to the intervention itself, which combine and interact. The mechanisms embedded in the social and organisational context can support or inhibit change. Change is not linear, and the outcome of an intervention can be change (morphogenesis) or no change (morphostasis).

**Figure 2:**
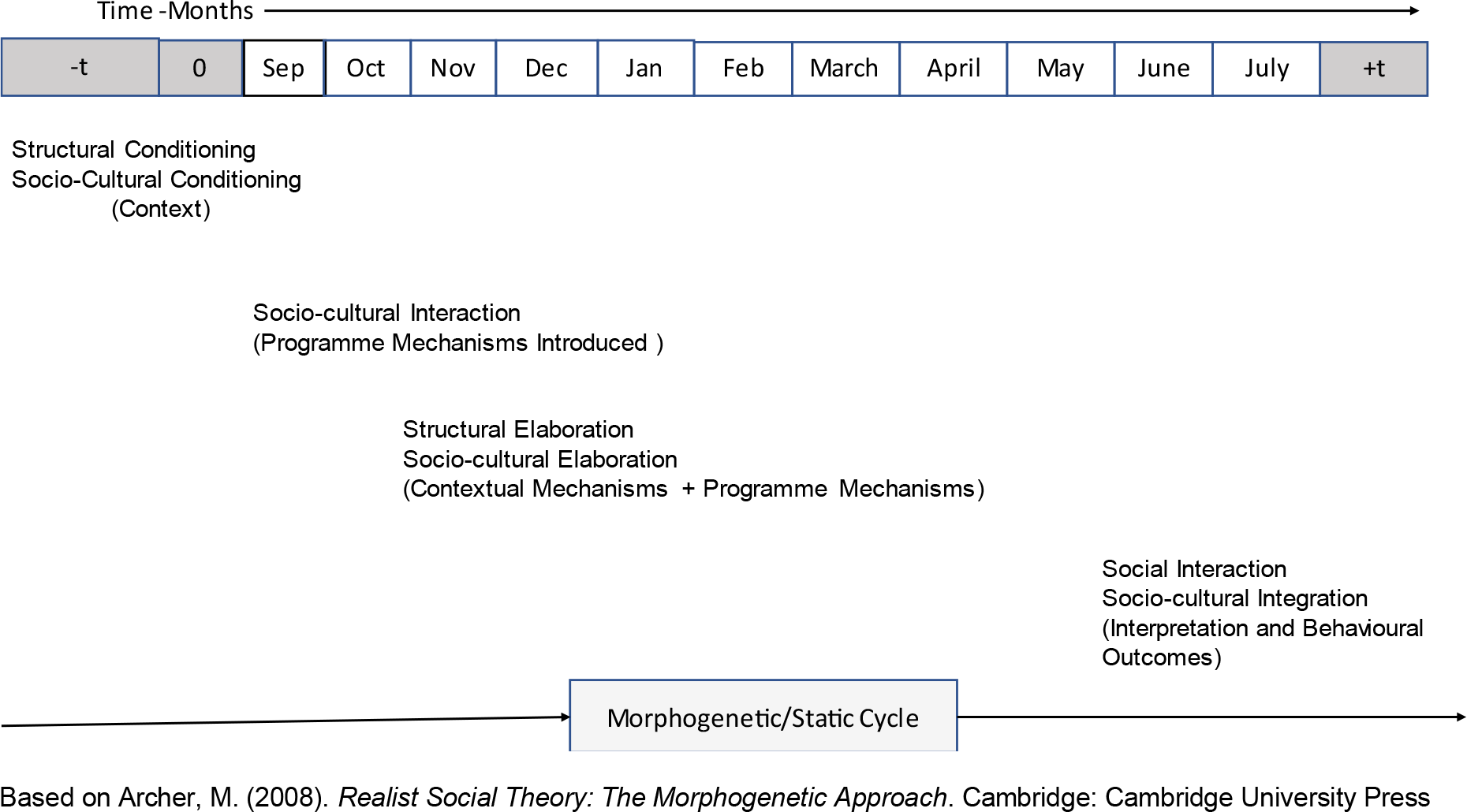
Intervention Cycle Showing the CAIMO Configuration.

### Programme Theory

Our starting point is a conceptual model based on a rapid review of the literature on school mindfulness interventions to identify mechanisms (an ensemble of structures, processes, and relations) that could be activated and bring about change (Figure 3). However, most research has been carried out by psychologists and has reported the individual-level outcomes rather than considering how the intervention brought about the changes. Little attention has been paid to how contexts change when CAs and their teachers practice mindfulness. Evidence, for example, suggests that group effects are important (77), as indicated by findings from whole-school interventions with mindful schools cultivating mindfulness as a disposition that changes pupil (and teacher) behaviour (44,56,63,68,78–82), thereby improving the school environment (56,83,84). A positive school climate is essential for positive student outcomes, including mental wellbeing, with feedback loops between teachers’ improved mental wellbeing and pupils’ (79,84–88).

**Figure 3:**
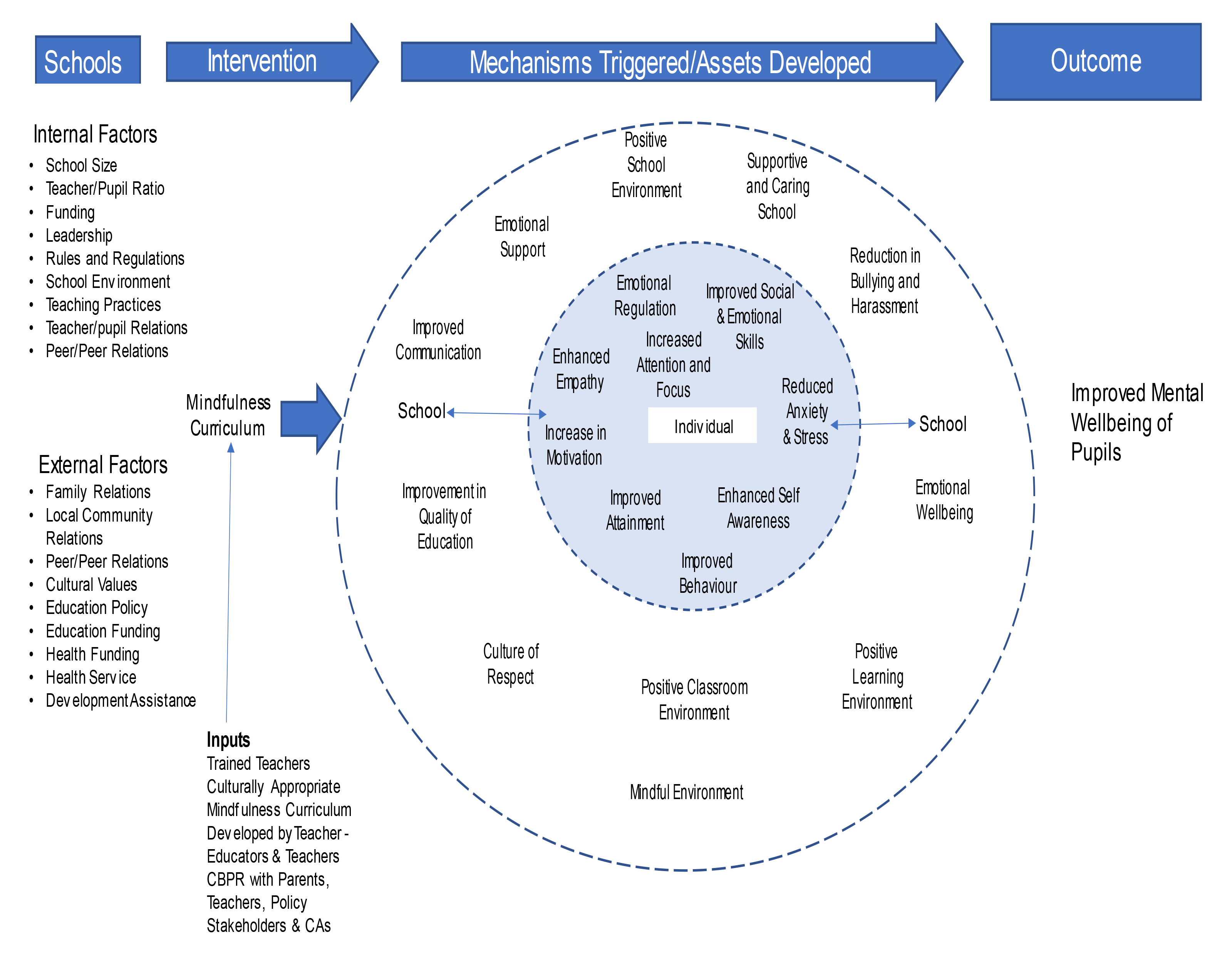
Programme Theory.

We hypothesise that mindfulness training will have a positive impact on CAs’ mental wellbeing through triggering mechanisms that increase the ability of CAs and their teachers to cope with adversity and improve the school climate and that this will provide a context in which CAs can improve their school performance and enjoy an improved subjective quality of life. The complex interaction and reinforcement through feedback loops between the context, CAs agency and these mechanisms will promote CAs’ mental wellbeing (Figure 3).

### Recruitment Process and Sampling of Schools

The research will be conducted in schools in Rwanda and Ethiopia, a 9-year basic school in rural Rwanda (7-16 years) and an urban primary school (7-14 years) in a deprived neighbourhood in Addis Ababa, Ethiopia. Three primary schools will be sampled in each country: a feasibility school for piloting the data collection tools and delivering mindfulness training, an intervention school, and a control/waitlist school. The inclusion criteria for schools in Ethiopia and Rwanda is that they are public schools or publicly maintained schools. In Ethiopia, the schools will be primary schools (pupils aged 7-15 years) in a deprived neighbourhood in Addis Ababa (Addis Ketema), and in Rwanda, they will be 9-year basic schools (7-16 years) in a rural area in Butaro district. Schools will be identified from a list of eligible schools falling in the interquartile range for school size (number of pupils) in the chosen districts in each country. Information on the number of older pupils relative to younger pupils per school will then be used to identify suitable interventions and control schools to achieve a reasonable balance across schools of the ratio between older and younger pupils. When potential schools have been identified, we will check with the district/city director of education on school suitability in terms of perceived ability to deliver the intervention. The three schools in each country will then be randomised into one of three groups: pilot school, intervention school and control/waiting list school. We will then contact the schools and invite them to participate in the project.

All pupils attending the intervention schools in Ethiopia and Rwanda will be eligible to engage in mindfulness training, subject to parental and CA informed consent. All teachers will be involved in teaching mindfulness. However, only CAs in years 4-9 in Rwanda and 4-8 in Ethiopia will complete the baseline, process and end-of-line questionnaires and psychometric tests with all the school teaching staff. Younger children are excluded from completing questionnaires and taking psychometric tests for literacy reasons and their inability to read and comprehend self-report questionnaires and tests with minimum help. All CAs and teachers will be eligible to be included in the qualitative research (interviews and focus group discussions). All mothers/primary carers will be surveyed at baseline and end-of-line.

The schools will be recruited in May of year 1 (2023). On recruitment, the feasibility and trial schools will be invited to each nominate five members of staff (20 in total) to be mindfulness champions. The mindfulness champions will be involved in developing the mindfulness intervention and training the teachers in their schools to deliver it with the support of the teacher educators. The trial schools will be asked to facilitate the recruitment of a purposive sample of mothers/primary caregivers and of CAs to become members of reference groups. The mothers /primary caregivers will be sampled so that there are mothers in the group with children in classes across the school. The CAs will be selected so that there are equal numbers of boys and girls, and they come from classes across the school.

Incentives for the schools to take part include:

- £1,000 remuneration in recognition of administrative commitments;
- The opportunity to introduce a SBMI with support from experts in delivering SBMIs;
- The opportunity for all teachers to be trained in delivering a SBMI and selected teachers to be involved in the development of the intervention;
- Teachers to be financially compensated for the time spent training and the additional administrative work of delivering the SBMI.

### Sample Size

Power calculation - Building on the feasibility study and trial conducted in Uganda (61), using β .80; α .05; .30 s m s o 350 ou r s r qu r . A jus or 30% study attrition, the target final sample size to be recruited in each country is n=460 (230 children in each group). The number of 230 participants per school is lower than the average school roll in years 4-9 in rural Rwanda and 4-8 in Addis Ababa, Ethiopia, where there are approximately 400 pupils per primary school in both countries. The sample size will therefore be large enough to explore additional questions related to the critical realist evaluation (e.g., are there different outcomes according to gender, age, ability, class/teacher, school attendance, and socioeconomic circumstances).

## Research Implementation

The research will be carried out in two Phases. Figure 1 shows the phases of the implementation of the study, and Figure 3 shows how the data sources feed into the critical realist framework connecting context, agency, intervention mechanisms, and outcomes.

### Phase 1: developing and refining programme theory and the mindfulness intervention

In phase 1, as well as developing the mindfulness intervention, we will gather information from several sources to refine our programme theory (Figure 3 and Table 2). This will include two realist syntheses of the literature, one on mindfulness-based interventions in schools (89,90) and one on interventions to promote CAs’ mental wellbeing by improving the school environment in low-and middle-income countries (91). We will extract potential contextual factors, agency, programme mechanisms, and outcomes (CAIMO configuration) relating to how school mindfulness interventions promote CAs’ mental wellbeing. We will also do a critical realist scoping review of the literature that theorises how mindfulness works.

**Table 2:** Critical Realist Terminology and Definitions.

| Terminology | Definition |
| --- | --- |
| Complexity | The social world is an open system with constellations of structures and mechanisms that interact to produce outcomes. |
| Context | Contingent combination of social relations in which mechanisms are embedded. Context is relational and dynamic. It enables and constrains agency in response to an intervention, with the outcome being an unchanged or transformed context, morphostasis or morphogenesis. |
| Culture | The shared beliefs, values, and symbols shape how individuals understand and interpret the world around them. |
| Human Agency | The responses of the stakeholder (teachers and CAs) to the intervention |
| Intervention Mechanisms | Mechanisms that are embedded in the intervention with the aim of CAs using them to trigger new mechanisms that will promote their mental wellbeing. |
| Judgemental Rationality | Weighing up what is likely to be true in a given context – compare competing explanations and decide which one is most likely to be true |
| Laminated System | Laminated systems are characterised by emergent properties which arise from the interaction between the different levels of the system, chemical, biological, psychological, psychosocial, behaviours, social, cultural, and economic and cannot be reduced to the properties of the individual levels. The social system is complex and dynamic; the different layers are not independent but interconnected and influence each other in complex ways. Mechanisms at a lower level can create the conditions for unfolding mechanisms at a higher level that can combine and create something new. |
| Mechanisms | Mechanisms cause something to happen; they generate observable events. They are not directly observed but inferred from their effects. It is necessary to differentiate between context mechanisms and intervention mechanisms. Mechanisms belong to different strata of reality, physical, biological, psychological, psychosocial, socioeconomic, cultural, and normative. |
| Structure | The concrete and material aspects of social organisation, such as institutions, organisations, and social networks |

An initial political economy analysis (PEA) will review academic and grey literature to document the context in Ethiopia and Rwanda, including health and educational policies relating to CAs, the health and educational status of CAs, and the community and school context within which CAs live their daily lives. We will also undertake a stakeholder analysis to identify policy stakeholders to interview. A purposive sample of these stakeholders will include Rwandan political and policy organisations and development partners (officials and NGOs) representatives active in the education and health sectors and city-level organisations in Ethiopia. In-depth, semi-structured interviews and a workshop with policy stakeholders will augment the PEA—policy by exploring policy, services, and economic influences on CAs’ mental wellbeing.

Participatory action research (PAR) will also be progressed in Phase 1, with mothers/primary caregivers and CAs, to empower them to have a say in the intervention design. After that, participants (parents and CAs) will be invited and supported to adopt parallel 'whole systems' research roles extending codesign of the intervention into implementation, evaluation, and dissemination through reference groups (92). The reference groups will form the sample for the parents and CAs PAR. PAR will elicit and systematise lived experience on the structural and cultural influences on CAs’ mental wellbeing and the cultural factors that the mindfulness intervention needs to take account of and appraise and adapt the draft mindfulness intervention.

Baseline questionnaires, interviews and focus group discussions (FGDs) for the main carer of CAs at the intervention schools and their teachers will also provide contextual information on carer and teacher mental wellbeing, the school 'climate', school-community relations, poverty, and deprivation The teacher-educator interviews will explore how they think the programme will work and what mechanisms they have identified that may block or inhibit the mindfulness intervention.

The research tools will be drafted in English and translated into local languages by native speakers. Local language translation will be quality assured by two native speakers working independently. If necessary, any disagreements will be resolved by bringing in a third native speaker. In phase 1, the PAR will be carried out in local languages. Policy stakeholder interviews, workshops, and teacher-educator interviews will be in English or the local language, depending on participants’ preferences. Policy stakeholder workshops will be in English.

Achieving meaningful outputs will require sophisticated teamwork and pragmatic approaches. We will use rapid qualitative research methods (QRM) to combine the depth of qualitative research with speed of data collection and analysis. Rapid QRM is a fast-developing area to study and evaluate services and interventions. Rapid QRM is participatory, iterative, problem-driven, highly team-based, with data collection and analysis occurring in parallel. We will draw on methods and standards Rapid Research Evaluation and Appraisal Lab (RREAL) developed. The RREAL Sheet process supports the systematic collection of data according to the main research questions, a rapid summary of findings and identification and triangulation of emerging themes, supporting the identification of convergences, divergences, and gaps, including for further adaption of data collection and analytical processes through regular supportive team reflection (93)

Qualitative interviews with policy stakeholders and teacher educators will be transcribed verbatim and translated into English when necessary. Notes from workshops (taken independently by two notetakers) will be reviewed by the members of the research team facilitating the workshop and then transcribed. The data from the PAR, the policy stakeholder and teacher interviews will be coded in the CAIMO configuration. Findings from Phase 1 will be used to refine programme theory and inform the design of the mindfulness intervention.

### Phase 2: Testing the Programme Theory and Economic Analysis

In Phase 2, we will test our programme theory and conduct an economic (cost) analysis (Figures 1 and 4).

**Figure 4:**
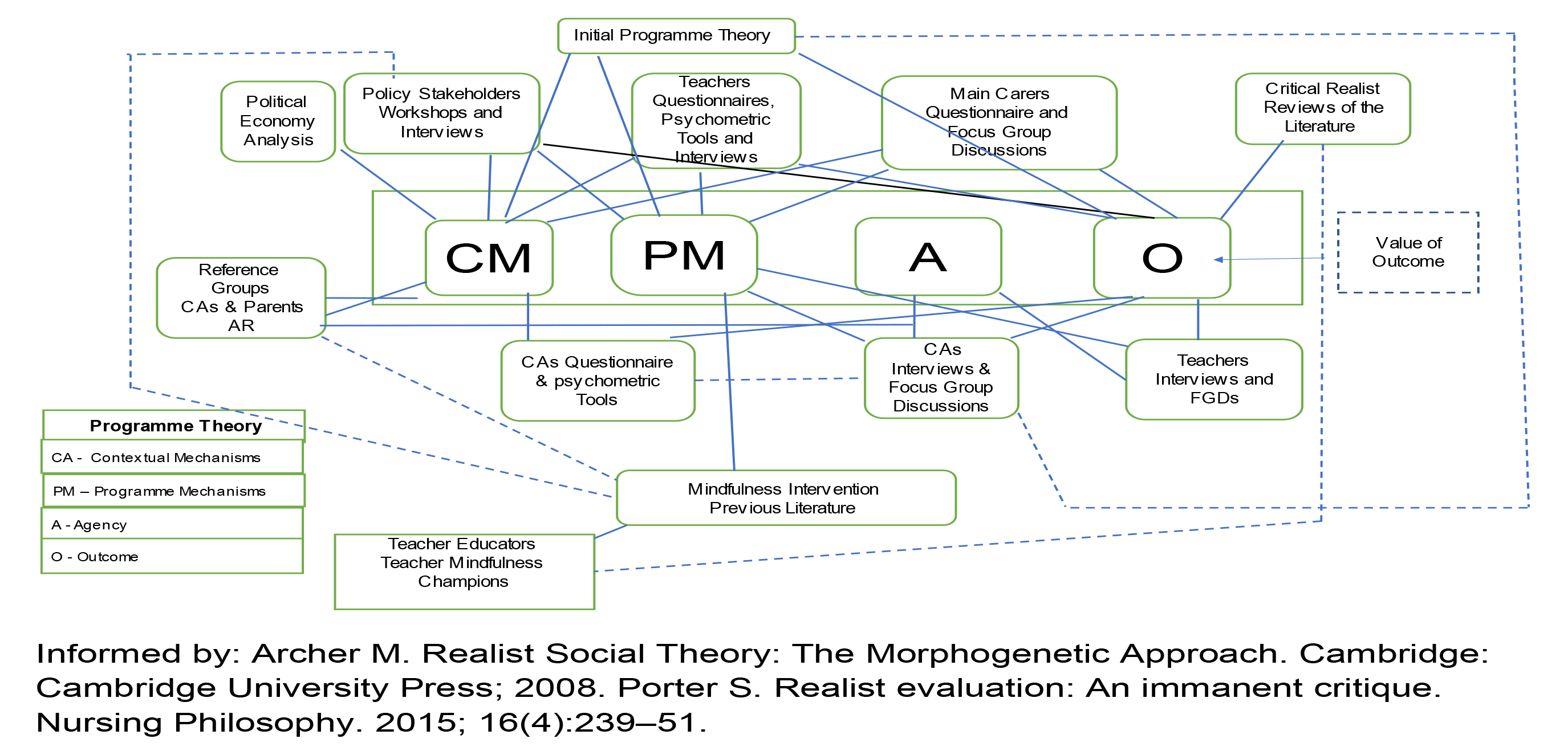
'Plumbing Diagram' Showing How the Data Sources Feed into the Critical Realist Framework Connecting Context, Mechanisms, and Outcomes.

#### The Implementation of the Trial

Table 3 shows the timeline for the trial activities. The trial will occur over one school year, from September 2024 to July 2025. Before implementation of the trial, a two-month pilot will be carried out to 'test' the use of the psychometric tools and the teaching of mindfulness in a pilot school in each country(-t1). The tools and teaching methods will be reviewed and revised as necessary before the start of the trial in the intervention schools. Data will be collected at baseline and close-out, with process data collected while the mindfulness curriculum is taught in the intervention school (t1, t2, t3).

**Table 3:** Methods for refining the initial programme theory before the Mindfulness programme is implemented in schools.

| Steps | Question | Methods | Outputs |
| --- | --- | --- | --- |
| Identification of the potential contextual factors | What are the conditions in Rwanda and Ethiopia that impact CA mental wellbeing? | Political economy analysis. Interviews/workshops/ PR with policy actors, parents, CAs and teacher educators. Baseline questionnaires – main carers & teachers. | Potential contextual factors (mechanisms) identified. |
| CR synthesis | What is the evidence on the effectiveness of mindfulness interventions in promoting CAs' mental wellbeing in LMICs? What are the mechanisms that are triggered? What programme theories have been tested with what outcomes? | CR review of mindfulness interventions for promoting CAs' mental wellbeing in LMICs, school climate interventions in LMICs with mental wellbeing as a primary or secondary outcome was CA mental wellbeing and theories of how mindfulness works to promote mental wellbeing. | Identifying the mechanisms through which the outcomes were brought about as well as the required contextual conditions. |
| Eliciting the programme theory of the developers | How is the mindfulness intervention supposed to work? What are the assumptions of the developers? | Interviews with the teacher educators and a review of the mindfulness training programme. | The assumptions of the actors on the mechanisms through which they believe the mindfulness intervention will promote CAs' mental wellbeing. |
| Refining the mechanisms | Revising the initial theories explaining how the mindfulness intervention is expected to work to promote CAs' mental wellbeing. | Review of concepts and theories | Refine mechanisms and potential middle-range theories. |

**Figure 5:**
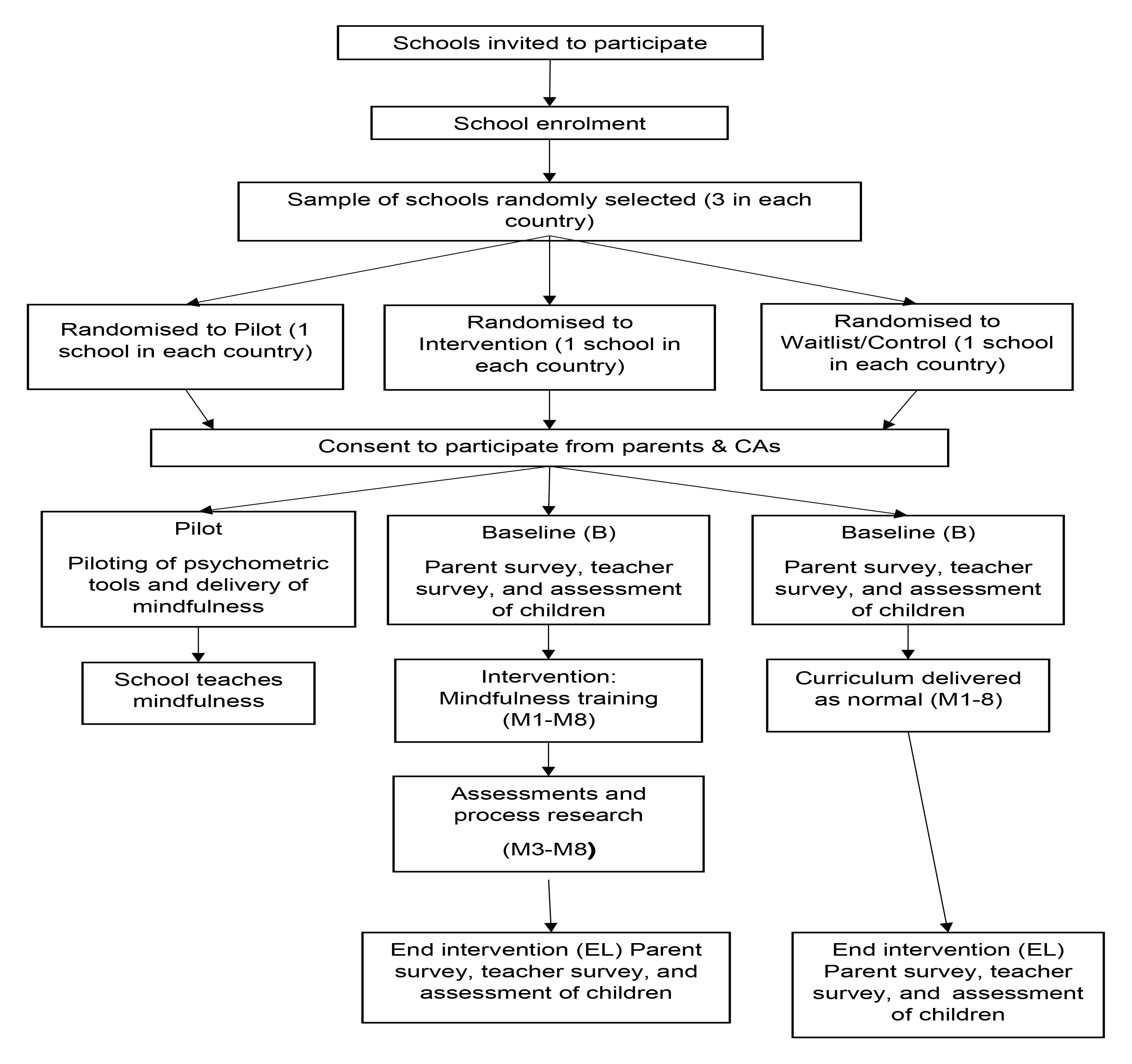
CONSORT Flow diagram of participants through the mindfulness trial.

#### Data Collection Methods

We take the stance that technical data collection methods should not be conflated with methodology, the logic of enquiry (76,94–96). Quantitative data, including psychometric tools, will provide numerical measurements and statistical analyses that will be useful for measuring the impact of the mindfulness intervention and patterns in the data that are indicative of underlying structures, mechanisms, or constraints (97). Qualitative data will enable us to understand the underlying mechanisms and processes that generate these patterns. Triangulation of quantitative and qualitative data is used for confirmation, completeness and abductive inspiration (98,99). Multiple methods and triangulation will support causal analysis based on different data types and sources, analytical methods and multiple investigators and theories (100).

Figure 4 and Table 4 map data collection methods and tools against the three main data collection points in Phase 2 and research participants. Additional Information 1 maps sources of evidence and examples of the questions we will ask to enable us to identify mechanisms to explain why the mindfulness intervention had the impact it did (101). We will collect quantitative data (administrative, survey, and psychometric data) and qualitative data (interviews, FGDs, PAR and observation). Quantitative methods will be used to collect data to answer what questions. Qualitative methods will ask explanatory, how and why questions, asking participants how they interpret the causal process (101,102). Participants will include CAs, mothers/main carers, teachers, school administrators and policy actors. Data will be collected before the mindfulness intervention is delivered for intervention and control schools (baseline data) and at the end of the school year in which the mindfulness intervention was delivered (end-of-line). In the intervention schools, qualitative and quantitative process data will be collected during the intervention.

**Table 4:**
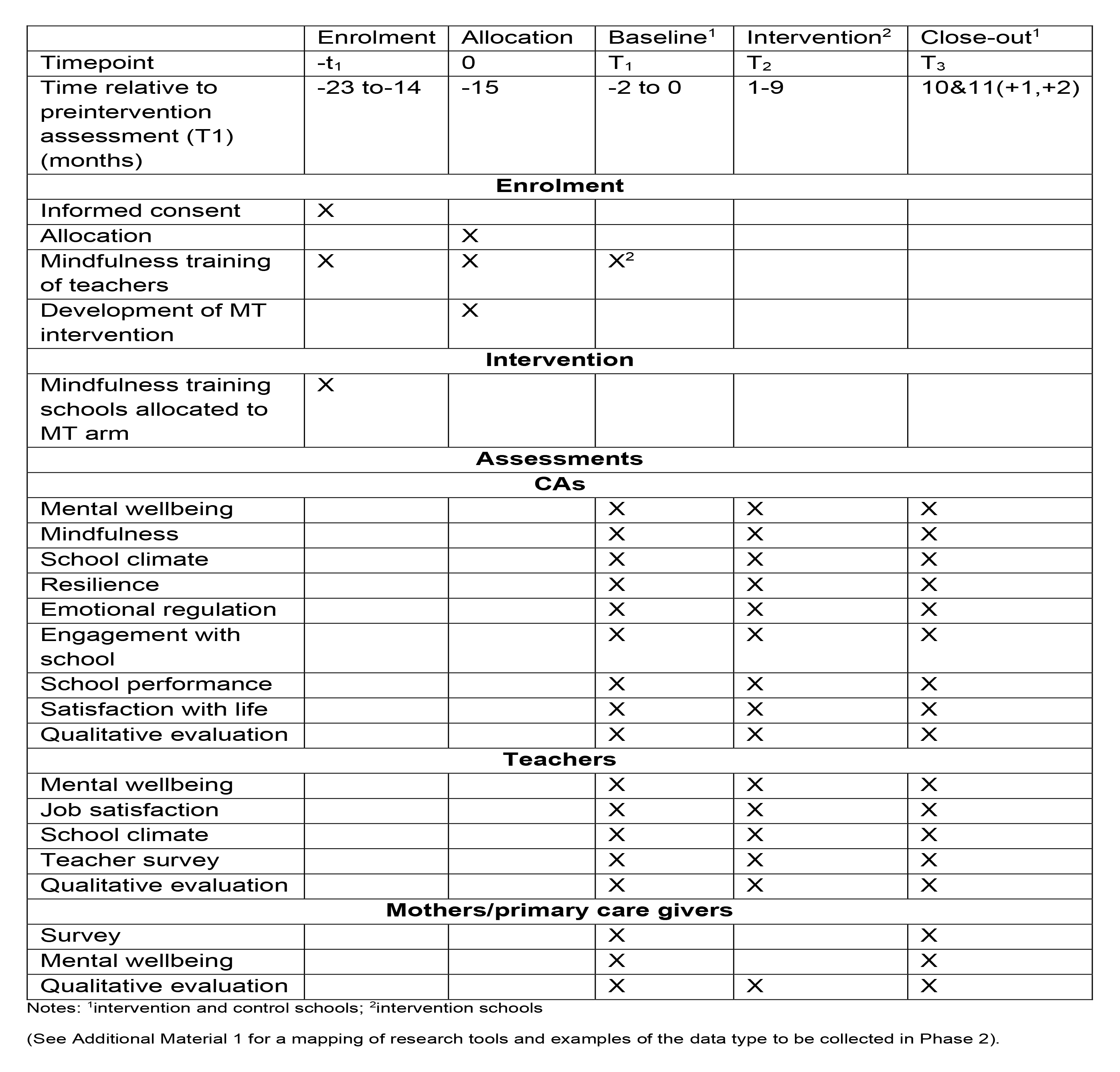
Standard Protocol Item: Recommended for Intervention Trials (Spirit) diagram detailing trial activities and measures and their timing.

The measures we will use to assess the impact of the mindfulness intervention will be finalised when we have completed the three literature reviews and refined our programme theory. The ones listed here are indicative.

The primary outcome of interest is CAs’ mental wellbeing.

##### Indicative main CAs outcome measure

Acholi Psychological Assessment Instrument (61) Indicative secondary outcome measures for children:

1. Quality of life: Cantril’s Ladder (103)
2. School Climate: Beyond Blue School Climate Survey (104)
3. Pupil Behaviour: In-school Behaviour Survey (children’s version) (105)
4. Mindfulness: Child and Adolescent Mindfulness Measure (106)

##### Indicative measures for teachers

1. Mental wellbeing: Kesler Psychological Destress Scale (107)
2. Job satisfaction: Teacher Job Satisfaction Questionnaire (108)
3. Quality of life: Cantril’s Ladder (103)
4. School climate: Beyond Blue School Climate Survey (104)

##### Indicative Measures for mothers/main carers

1. Mental wellbeing: Kesler Psychological Destress Scale (107)
2. Quality of life: Cantril’s Ladder (103)
3. Child Behaviour Questionnaire (parents’ version)(109)

All research tools will be translated into local languages by native speakers, reviewed by the reference groups, and revised as necessary. The local language versions will then be quality assured by two independent native speakers in each country to ensure that the tools in English and local languages convey the same meaning. Any disagreements will be discussed by the reviewers and resolved with the help of a third native speaker, if necessary. Psychometric tools will be translated into local languages by two independent translators in each country (apart from those for CAs in Rwanda, where pupils are taught in English from primary 1), and any disagreements will be resolved. If necessary, the psychometric tools will be validated for use in LICs. All tools will be piloted and revised in the light of feedback.

#### Quantitative Data Collection

All quantitative data collection will be carried out face-to-face by trained enumerators supervised by experienced researchers. The mothers/main carriers will be interviewed in their own homes and pupils and teachers at schools. All the mothers/main careers and teachers of children attending the intervention and control schools will be invited to participate in the questionnaire study. All CAs in grade four and above will be invited to participate. Teachers and CAs questionnaires will be self-completion with teachers assisted by research assistants, supervising completion by CAs. Mother/main carers surveys will be quality assured by a 10 per cent callback and a 10 per cent sampling of completed questionnaires.

#### Qualitative Data Collection

All qualitative research, except for policy stakeholders, will be conducted in local languages. The parents, CAs from the intervention schools, and policy stakeholders participating in Phase 1 PAR will be invited to participate in Phase 2 PAR. In addition, at the intervention schools: all the teachers will be invited to participate in semi-structured interviews between 16 and 20 in each school; two children from each class, one boy and one girl, between 32 and 40 in each school, will be invited to participate in interviews and FGDs; and 40 parents will be invited to participate in interviews or FGDs. The same teachers, pupils and parents will be invited to participate in the baseline, end-of-line, and process evaluation. Trained observers will carry out participant observation in classrooms and the school playgrounds using a qualitative tool to record the behaviour of CAs and teachers. Qualitative interviews and FGDs will be recorded, with participants’ permission, transcribed verbatim and translated into English. Notes will be taken at workshops and transcribed in English. The PAR (Phases 1 and 2) will adopt the RREAL Sheet process.

#### Data Analysis

The main aim of the data analysis is to explain how and why the mindfulness intervention had the impact it had on CAs’ mental wellbeing using the RRRIREI© Framework (Table 1). According to this framework, causation occurs when the agents’ reaction to an intervention in a given context triggers mechanisms that produce outcome(s). In analysing the data, we will move from the description of changes to a description of the process that conditioned the changes. We will then use abduction, retroduction and retrodiction to refine our programme theory. There are three stages to theory development (110). The first is the description of the observed empirical events. The second is the identification of the mechanisms triggered by agents in response to the mindfulness intervention and the structural, cultural, and context changes that took place using CAIMO configurations, retroduction and retrodiction to develop trans-factual theories integrating disciplinary explanations and elimination using judgemental rationality to eliminate the theories that lack explanatory power (7). The third is using judgemental rationality to identify the most comprehensive interdisciplinary explanation. This will be followed by developing change pathways (including those that are non-linear and characterised by emergence and feedback) showing how complex interactions of mechanisms (context mechanisms that predate the intervention and those triggered by the intervention) support change, as well as those that restrict or prevent change, leading to observed outcomes (111–114).

#### Quantitative Analysis

A statistical analysis plan will be agreed upon before database lockdown. Data will be recorded and analysed using standard statistical software packages (e.g., STATA, R, SPSS) in preparation for analysis, creating a record for every CA respondent in the intervention and control schools.

The quantitative analysis of the effectiveness of the intervention, for whom it was effective and under what circumstances will be measured by calculating the difference in the change in primary outcome measure score between intervention and control groups over time. The primary analyses will be on an intention-to-treat basis at the 9-month end point, adjusted for baseline values of the outcome measure and variables for which randomisation did not achieve a reasonable balance between the arms at baseline or those associated with missing outcome data. Analyses of primary and secondary outcomes will be conducted using Generalised Linear Modelling for continuous outcomes, with appropriate link and distribution functions applied (e.g., SDQ Total Difficulties score) and generalised (logistic) regression models for binary outcomes. Intervention effects will be presented as adjusted mean differences and effect sizes (ES), defined as standardised mean differences, with 95% confidence intervals (CIs) for continuous outcomes and adjusted odds ratios with 95% CIs for binary outcomes.

Factor and path analyses will draw out the latent variables and causal connections to support theory construction (97). This will include constructing a 2-1-1 multilevel mediation model that includes an intervention at country level and mediators and outcome measures at level 1, enabling the disaggregation of the impact mediator on the outcome at the country level (context) and CA level (115,116).

#### Qualitative Analysis

The qualitative analysis will explore what it is about the conditions in the intervention schools that enabled certain actions to occur and allowed certain consequences to follow (102,117,118). The qualitative data enables an in-depth, 'thick' exploration of key analytical constructs, i.e., agency and mechanisms, directly through participants’ accounts of how they think the mindfulness intervention worked and indirectly through their accounts of their experiences of related actions.

The qualitative data will be analysed using Template Analysis, a form of thematic analysis that uses a priori codes and identifies unanticipated themes (119). This allows pre-existing theory to guide the contextualisation and redescription of the data (7,73). The analysis will identify three types of codes: experiential (what observed experiences and events in the 'empirical domain'), inferential (why, the context mechanisms that generated the experiences) and dispositional (how, the outcomes) (110)—moving from what is observable (the empirical) to what is happening (the actual) to identifying the causal powers (the real). NVivo and Excel spreadsheets will be used to manage and analyse qualitative data.

The data will be coded using abductive reasoning, re-describing what is observed in terms of theories to reveal the possible mechanisms responsible for the observed outcomes. The data will then be mapped more precisely using the CAIMO configuration, context, agency, intervention, mechanisms, outcomes, and patterns in the data identified. Retroductive reasoning will be used to identify the causal mechanisms and the necessary conditions for these mechanisms to work, asking what the world must be like for the phenomena we observe to be as they are and not otherwise (102). Retrodictive reasoning will then be used to identify the combination of these mechanisms that best explain the empirical results.

#### Health Economics Analysis

The Cost-effectiveness of the mindfulness intervention will be assessed by measuring and valuing the amount of teacher training time, the time trainers take to deliver the training and any expenses associated with training manuals. Health care costs and household expenses (out-of-pocket) associated with managing mental health problems amongst pupils will also be measured. Costs data will then be combined with CHU9D data (or similar) to calculate Quality Adjusted Life Years (QALYs) and cost per QALY gained (DALYs may also be computed, subject to the use of alternative quality-of-life measures). Generalised linear modelling will be used to estimate the difference in costs and QALYs attributable to the intervention, summarising this by constructing Cost-Effectiveness Acceptability Curves. Appropriate sensitivity analysis will be conducted to test the robustness of study findings on cost-effectiveness. Finally, the cost of rolling out the intervention more widely to other settings will be estimated by modelling levels of resource use for the education system and associated costs which apply to new schools interested in take-up and delivery of the intervention.

## Ethical Considerations, Data Protection and Safeguarding

The project has been approved through the ethics review procedures at the University of Aberdeen, Addis Ababa University, and the University of Rwanda. We recognise that ethical and safeguarding considerations are essential to ensure the protection and wellbeing of the parents, CAs, bystanders and junior researchers involved in the research. The project has safeguarding and whistleblowing policies (120). All researchers, including enumerators and notetakers, will be trained in research ethics, governance, data management, safeguarding, and researching with children (121). All participants will be required to give written informed consent or a thumbprint, and the mothers of children under 18 years will be asked to consent to their child’s participation and the children themselves. All participants will be given a participant information sheet.

The Data Management Plan has been approved by the UOA’s Research Applications and Data Management team. It is designed to protect the integrity and security of data and the confidentiality of research participants. All data will be stored to prevent data linkage, and all linking data will be destroyed following the third round of data collection in year four. All data arising from the project will be logged in an open-access repository within six months of the completion of the project under Creative Commons. All data and other project records will be archived for five years following the completion of the project.

### Monitoring

The project will be monitored for quality, cultural appropriateness, and regulatory compliance. An independent advisory committee composed of academics, government officials, non-government organisations and donor agencies will participate in the monitoring of the project. The Management Committee will review and update the Risk Register every three months.

## Community and Policy Stakeholders Involvement

The project takes a Multi-layered Stakeholder Input Approach. Parents, CAs, teachers and policy stakeholders are involved at every stage of the research through involvement in the development and design of the mindfulness intervention, the implementation of the trial and the interpretation of the findings. The stakeholder analysis will enable us to ensure that we communicate with those most interested in the research findings and those with the most significant influence over policy.

A sample of CAs and mothers/primary carer givers from the intervention schools will be selected to become members of reference groups. The reference group members will meet regularly throughout the project and discuss in years 1 and 2 what they think are the structural and cultural influences on CA mental wellbeing, the cultural factors that the mindfulness intervention needs to consider, and comment on the draft mindfulness intervention. In years 3 and 4, they will discuss what changes have happened at school, at home and in the community since the mindfulness programme was implemented and why they think these changes have occurred. In year 4, they will be asked to comment on the theories that have been developed. The teacher mindfulness champions will be involved in designing and delivering the intervention. All teachers in the intervention school will teach the mindfulness curriculum, and a purposive sample will be asked to participate in interviews and FGDs. They, too, will be asked about changes in the children and themselves since the mindfulness intervention was introduced. There will be observations of the school and classrooms in the intervention schools at baseline, during delivery and at the end-of-line.

## Communication and Dissemination of Results

A communications and publication strategy has been developed to ensure the dissemination of knowledge arising from the project to the broadest possible audience (122). Findings from the project will be disseminated ina variety of outlets and formats, including peer review journal articles, presentations, policy briefs and engagement with political actors and disseminated via academic publications, webinars, social media, and the project website.

We are targeting four main academic audiences, heath scholars with interest in CAs’ health and wellbeing, education scholars with interest in enabling CAs to develop to their full potential, development studies academics with interest in development and the wellbeing and education of CAs, and scholars with interest in using CR in empirical research for policy and practice. We are targeting three main policy actor audiences, those working in the health and education sectors in Rwanda and Ethiopia, including Official Development Partners and NGOs, and regional and international/global organisations that are involved in the education and health sectors, including WHO, UNICEF, UNESCO, and the African Union. We also plan to disseminate the findings to the CAs, their parents, and teachers in the schools where we conduct the research to the public, more generally in Rwanda and globally. We will engage with the print and broadcast media to do the latter and publish articles in outlets such as The Conversation. All training materials, anonymised data sets and publications arising from the project will be published under a Creative Commons Licence, Creative Commons Attribution 4.0 Unported (CC BY 4.0) and available on the project web site: Open Access Training | Education | The University of Aberdeen (abdn.ac.uk).

## Study Governance

Pamela Abbott (PA) will direct the study with Wenceslas Nzabalirwa (WN) as Joint-Lead. PA will manage the team at the University of Aberdeen, WN in Rwanda, and Mahlet Yard and Tsion Hailu in Ethiopia. The intervention and research teams will be functionally independent. The research team for the trial will be managed by Paul McNamee, Elias Haile, and Eric Remera. PA, Lucia D’Ambruoso, Darius Gishoma, and Kibur Engdawork will manage the qualitative research and PAR. The intervention team will be managed by Graeme Nixon, Henok Hailu, and Ali Kaleeba Bakali.

## Conclusion

The findings from the research will provide teachers, policymakers and researchers with a deeper understanding of how and why mindfulness interventions work (or do not work) to promote the mental wellbeing of CAs. Combining a trial with critical realist evaluation will bring together the strengths of both approaches, providing a more comprehensive and nuanced understanding of the effects of the mindfulness intervention and the underlying mechanisms and contextual (social, cultural, and organisational) factors that shape those effects. The active involvement of stakeholders, including CAs, their main carers, teachers and policymakers, will enhance the evaluation’s relevance, validity, and applicability. Stakeholders can provide valuable insights into the contextual factors and mechanisms at play.

### Ethics approval and consent to participate

The ethics committees of the University of Aberdeen, UK, Addis Ababa University, Ethiopia and the University of Rwanda, Rwanda, have given ethics approval.

### Consent for publication

Not required

### Availability of data and materials

No data was used in preparing this protocol. The data sets that the project generates will be deposited with the UK National Data Archive within six months of the completion of the project under a Creative Commons Attribution 4.0 Unported (CC BY 4.0) license. The training materials produced by the project will be made available under a Creative Commons Attribution 4.0 Unported (CC BY 4.0) license on the project website: https://www.abdn.ac.uk/education/research/cgd/nihr-camw-subsaharan-africa/index.php

## Funding

The research is funded by the National Institute for Health and Care Research (NIHR133712) using UK aid from the UK Government to support global health research.

### Authors’ contributions

PA and LD conceived the research, and they, together with MY, TH, WN and PM, made substantial contributions to the conception of the project and the acquisition of funding. PA drafted the protocol, all authors reviewed it critically and made intellectual contributions, and PA drafted the final version. All authors have read and approved the manuscript and agree to be accountable for its content.

Pamela Abbott https://orcid.org/0000-0002-5013-343X

Lucia D’Ambruoso https://orcid.org/0000-0002-8505-3368

Mahlet Yared https://orcid.org/0000-0002-7258-379X

Paul McNamee https://orcid.org/0000-0002-4540-8718

Tsion Hailu https://orcid.org/0000-0001-7415-7797

Wenceslas Nzabalirwa https://orcid.org/0000-0002-8299-2001

### Disclaimer

The views expressed in this protocol are the views of the authors alone. They do not necessarily represent the views of the National Institute for Health Research, the Court of the University of Aberdeen, the Board of Directors of the University of Rwanda, or the Board of Addis Ababa University.

### Open access

This is an open-access article distributed in accordance with the Creative Commons Attribution 4.0 Unported (CC BY 4.0) license, which permits others to copy, redistribute, remix, transform and build upon this work for any purpose, provided the original work is properly cited, a link to the licence is given, and an indication of whether changes were made. See: https://creativecommons.org/licenses/by/4.0/.

## Data Availability

The data sets that the project generates will be deposited with the UK National Data Archive within six months of the completion of the project under a Creative Commons Attribution 4.0 Unported (CC BY 4.0) license.

https://www.abdn.ac.uk/education/research/cgd/nihr-camw-subsaharan-africa/index.php

## References

1. About the Project [Internet]. Centre for Global Development. 2023 [cited 2023 May 8]. Available from: https://www.abdn.ac.uk/education/research/cgd/nihr-camw-subsaharan-africa/about-the-project-1616.php

2. Mindfulness in Schools Project. Teach Paws b [Internet]. Mindfulness in Education. 2023 [cited 2023 Mar 27]. Available from: https://mindfulnessinschools.org/teach-paws-b/

3. Porter S, McConnell T, Clarke M, Kirkwood J, Hughes N, Graham-Wisener L, et al. A critical realist evaluation of a music therapy intervention in palliative care. BMC Palliat Care. 2017;16(1):1–12.

4. Porter S, McConnell T, Reid J. The Possibility of Critical Realist Randomised Controlled Trials. Trials. 2017;18:133.

5. Porter S, O’Halloran P. The use and limitaion of realistic evaluation as a tool for evidence-based practice: A critical realist perspective. Nurs Inq. 2012;19(1):18–28.

6. Porter S. The uncritical realism of realist evaluation. Evaluation. 2015;21(1):65–82.

7. Danermark B, Ekström M, Karlsson JC. Explaining Society: Critical Realism in the Social Sciences. London and New York, NY: Routledge; 2019.

8. WHO. Adolescent mental health [Internet]. Fact Sheet. Geneva: World Health Organisation; 2022 [cited 2022 Dec 4]. Available from: https://www.who.int/news-room/fact-sheets/detail/adolescent-mental-health

9. Orth Z, Wyk B van. Adolescent Mental Wellness: A Systematic Review Protocol of Instruments Measuring General Mental Health and Wellbeing. BMJ Open. 2020;10(e037237).

10. Cortina MA, Sodha A, Fazel M, Ramchandani PG. Prevalence of Child Mental Health Problems in Sub-Saharan Africa: A Systematic Review. Vol. 166, Archives of Pediatrics and Adolescent Medicine. American Medical Association; 2012. p. 276–81.

11. Jorns-Presentati A, Napp AK, Dessauvagie AS, Stein DJ, Jonker D, Breet E, et al. The prevalence of mental health problems in sub-Saharan adolescents: A systematic review. PLoS One. 2021;16(5 May):1–23.

12. Sequeira M, Singh S, Fernandes L, Gaikwad L, Gupta D, Chibanda D, et al. Adolescent Health Series: The status of adolescent mental health research, practice and policy in sub-Saharan Africa: A narrative review. Trop Med Int Heal. 2022;27(9):758–66.

13. Mbwayo A, Kumar M, Mathai M, Mutavi T, Nungari J, Gathara R, et al. Strengthening System and Implementation Research Capacity for Child Mental Health and Family Wellbeing in Sub-Saharan Africa. Glob Soc Welf. 2021;

14. Juma K, Wekesah FM, Kabiru CW, Izugbara CO. Burden, Drivers, and Impacts of Poor Mental Health in Young People of West and Central Africa: Implications for Research and Programming. In: McLean ML, editor. West African Youth Challenges and Opportunity Pathways, Gender and Cultural Studies in Africa and the Diaspora. Cham: Palgrave MacMillan; 2020.

15. WHO. Mental health of adolescents [Internet]. World Health Organisation. 2021 [cited 2023 Mar 27]. Available from: https://www.who.int/news-room/fact-sheets/detail/adolescent-mental-health

16. Wickersham A, Dickson H, Jones R, Pritchard M, Stewart R, Ford T, et al. Educational attainment trajectories among children and adolescents with depression, and the role of sociodemographic characteristics: Longitudinal data-linkage study. Br J Psychiatry. 2021;218(3):151–7.

17. Pozuelo JR, Desborough L, Stein A, Cipriani A. Systematic Review and Meta-analysis: Depressive Symptoms and Risky Behaviors Among Adolescents in Low-and Middle-Income Countries. J Am Acad Child Adolesc Psyyciatry [Internet]. 2022;61(2):2550276. Available from: https://www.sciencedirect.com/science/article/abs/pii/S0890856721003105

18. Clark H, Coll-Seck AM, Banerjee A, Peterson S, Dalglish SL, Ameratunga S, et al. A Future for the World’s Children? A WHO–UNICEF–Lancet Commission. Lancet [Internet]. 2020;395:605–58. Available from: https://www.thelancet.com/article/S0140-6736(19)32540-1/fulltext

19. Kutcher S, Perkins K, Gilberds H, Udedi M, Ubuguyu O, Njau T, et al. Creating Evidence-Based Youth Mental Health Policy in Sub-Saharan Africa: A Description of the Integrated Approach to Addressing the Issue of Youth Depression in Malawi and Tanzania. Front Psychiatry. 2019 Aug;10:542.

20. Atilola O. Child Mental-health Policy Development in sub-Saharan Africa: Broadening the Perspectives using Bronfenbrenner’s Ecological Model. Health Promot Int. 2014 Apr;32(2):380–91.

21. Silwal AR, Engilbertsdottir S, Cuesta J, Newhouse D, Stewart D. Poverty and Equity. Global Estimate of Children in Monetary Poverty: An Update [Internet]. Washington DC: World Bank; 2020. Available from: https://openknowledge.worldbank.org/bitstream/handle/10986/34704/Global-Estimate-of-Children-in-Monetary-Poverty-An-Update.pdf?sequence=1&isAllowed=y

22. Trines S. Education in Ethiopia [Internet]. Education Systems Profiles. 2018 [cited 2022 Nov 5]. Available from: https://wenr.wes.org/2018/11/education-in-ethiopia

23. Trines S. Education in Rwanda [Internet]. Education System Profiles. 2019 [cited 2022 Nov 5]. Available from: https://wenr.wes.org/2019/10/education-in-rwanda

24. Pontalti K. School Violence: A Global Preventable Epidemic. Kigali: Plan Rwanda; 2013.

25. Murphy M, Jones N, Yadete W, Baird S. Gender-norms, violence and adolescence: Exploring how gender norms are associated with experiences of childhood violence among young adolescents in Ethiopia. Glob Public Health [Internet]. 2021;16(6):842–55. Available from: https://doi.org/10.1080/17441692.2020.1801788

26. The African Child Policy Forum, Save the Children Sweden. Violence Against Children in Ethiopia: In Their Words. 2006.

27. Ministry of Health. Violence Against Children and Youth Survey. Kigali: Republic of Rwanda; 2017.

28. Chuta N, Morrow V, Pankhurst A, Pells K. Understanding Violence Affecting Children in Ethiopia: a Qualitative Study. Young Lives Working Paper 188 [Internet]. Oxford: Oxford Department of International Development, University of Oxford; 2019. Available from: https://www.younglives.org.uk/sites/www.younglives.org.uk/files/YL-WP188 revised_0.pdf

29. Rwanda Biomedical Centre. Rwanda Mental Health Survey. Kigali: Rwanda Biomedical Centre; 2018.

30. Ministry of Health. National Mental Health Strategy 2012/13 - 2015/16. Addis Ababa: Federal Democratic Republic of Ethiopia Ministry of Health; 2012.

31. Ministry of Health. National Adolescent and Youth Health Strategy. Vol. 1. Addis Ababa: Federal Republic of Ethiopia; 2016. p. 1–100.

32. Ndagijimana A. Adolescent Mental Health Landscape Assessment in Rwanda Final Report [Internet]. Kigali: UNICEF, Rwanda Country Office; 2020. Available from: https://www.unicef.org/rwanda/reports/adolescent-mental-health-landscape-assessment-rwanda

33. Khodr A. Taking stock of children’s mental and psychosocial wellbeing in Ethiopia during COVID-19 [Internet]. UNICEF Ethiopia. 2020 [cited 2022 Nov 4]. Available from: https://www.unicef.org/ethiopia/stories/taking-stock-childrens-mental-and-psychosocial-wellbeing-ethiopia-during-covid-19

34. Ministry of Health. National Mental Health Strategy 2019-25. Addis Ababa: Federal Ministry of Health; 2019.

35. Ministry of Education. National School Health Policy. Kigali: Republic of Rwanda; 2014. p. 1–44.

36. Ministry of Education. National School Health Strategic Plan. Kigali: Republic of Rwanda; 2014.

37. Elizabeth P, Dutton R, Jones N, Baird S, Woldehanna T. Adolescent Psychosocial Wellbeing in Ethiopia: Implications for Policy and Programming from the GAGE Midline Data. Vol. 2020. London: Gender and Adolescent Global Evidence; 2021.

38. Deighton J, Lereya ST, Morgan E, Breedvelt J, Martin K, Feltham A, et al. Measuring and Monitoring Children and Young People’s Mental Wellbeing: A Toolkit for Schools and Colleges [Internet]. London: Public Health England and Anna Freud National Centre for Children and Families; Available from: https://www.annafreud.org/media/4612/mwb-toolki-final-draft-4.pdf

39. Bryant G, Heard H, Watson J. Measuring Mental Wellbeing in Children and Young People [Internet]. London: Public Health England; 2015. Available from: https://assets.publishing.service.gov.uk/government/uploads/system/uploads/attachment_data/file/768983/Measuring_mental_wellbeing_in_children_and_young_people.pdf

40. Goldberg JM, Sklad A, Elfrink TR, Schreurs KMG, Bohlmeijer ET, Clarke AM. Effectiveness of Interventions Adopting a Whole School Approach to Enhancing Social and Emotional Development: A Meta-analysis. Eur J Psychol Educ. 2019;34.

41. Caldwell DM, Davies SR, Hetrick SE, Palmer JC, Caro P, José L-L, et al. School-based Interventions to Prevent Anxiety and Depression in Children and Young People: A Systematic Review and Network Meta-analysis. Lancet Psychiatry. 2019;6:1011–20.

42. ŠouláKová B, Kasa A, Butzer B, Winker P. Meta-Review on the Effectiveness of Classroom-Based Psychological Interventions Aimed at Improving Student Mental Health and Wellbeing, and Preventing Mental Illness. J Prim Prev. 2019;40:255–78.

43. Phan ML, Renshaw TL, Caramanico J, Greeson JM, MacKenzie E, Atkinson-Diaz Z, et al. Mindfulness-Based School Interventions: a Systematic Review of Outcome Evidence Quality by Study Design. Mindfulness (N Y). 2022;13(7):1591–613.

44. Roeser RW, Mashburn AJ, Skinner EA, Choles JR, Taylor C, Rickert NP, et al. Mindfulness Training Improves Middle School Teachers’ Occupational Health, Wellbeing, and Interactions With Students in Their Most Stressful Classrooms. J Educ Psychol. 2022;114(2):408–25.

45. WHO. Helping Adolescents Thrive: Guidelines on Mental Health Promotion and Prevention Interventions for Adolescents. Geneva: World Health Organisation; 2020.

46. Panter-Brick C, Leckman JF. Resilience in Child Development – Interconnected Pathways to Wellbeing. J Child Psychol Psychiatry. 2013;54(4):333–336.

47. Kuyken W, Nuthall E, Byford S, Crane C, Dalgleish T, Ford T, et al. The effectiveness and cost-effectiveness of a mindfulness training programme in schools compared with normal school provision (MYRIAD): Study protocol for a randomised controlled trial. Trials. 2017;18(1):1–17.

48. Kapungu C, Petroni S, Allen NB, Brumana L, Collins PY, Silva M De, et al. Gendered Influences on Adolescent Mental Health in Low-income and middle-income Countries: Recommendations from an Expert Convening. Lancet Child Adolesc Heal. 2018;2:85– 6.

49. Berger R, Benatov J, Cuadros R, VanNattan J, Gelkopf M. Enhancing Resiliency and Promoting Prosocial Behaviour among Tanzan Primary-school Students: A School Based Intervention. Transcult Psychiatry. 2018;55(6):821–45.

50. Pillay K, Eagle G. The Case for Mindfulness Interventions for Traumatic Stress in High Violence, Low Resource Settings. Curr Psychol. 2019;

51. Langer ÁI, Medeiros S, Valdés-Sánchez N, Brito R, Steinebach C, Cid-Parra C, et al. A qualitative study of a mindfulness-based intervention in educational contexts in Chile: An approach based on adolescents’ voices. Int J Environ Res Public Health. 2020;17(18):1–17.

52. Barry MM, Clarke AM, Jenkins R, Patel V. A systematic review of the effectiveness of mental health promotion interventions for young people in low and middle income countries. BMC Public Health. 2013 Dec;13(1):835.

53. Bradshaw M, Gericke H, Coetzee BJ, Stallard P, Human S, Loades M. Universal School-based Mental Health Programmes in Low-and Middle-income Countries: A Systematic Review and Narrative Synthesis. Vol. 143, Preventive Medicine. Academic Press Inc.; 2021. p. 106317.

54. Hudson KG, Lawton R, Hugh-Jones S. Factors affecting the implementation of a whole school mindfulness program: A qualitative study using the consolidated framework for implementation research. BMC Health Serv Res. 2020;20(1):1–13.

55. Klingbeil DA, Renshaw TL, Willenbrink JB, Copek RA, Chan KT, Haddock A, et al. Mindfulness-based Interventions with Youth: A Comprehensive Meta-analysis of Group-design Studies. J Sch Psychol. 2917;63:77–103.

56. Weare K, Bethune A. Implementing Mindfulness in Schools: An Evidence-Based Guide [Internet]. London: The Mindfulness Initiative Sheffield; 2021. Available from: www.themindfulnessinitiative.org/appeal/donate

57. Krame K. The Efficacy of Mindfulness-Based Interventions on Occupational Stress and Burnout among K-12 Educators: A Review of the Literature. Vienna: World Conference on Teaching and Education; 2021.

58. Zarate K, Maggin DM, Passmore A. Meta-analysis of mindfulness training on teacher wellbeing. Psychol Sch. 2019;56(10):1700–15.

59. Klingbeil DA, Renshaw TL. Mindfulness-based interventions for teachers: A meta-analysis of the emerging evidence base. Sch Psychol Q. 2018;33(4):501–11.

60. Cook-Cottone C, Giambrone C, Klein J. Yoga for Kenyan Children: Concept-mapping with Multidimensional Scaling and Hierarchical Cluster Analysis. Int J Sch Educ Psychol. 2018;6(3):151–64.

61. Matsuba MK, Schonert-Reichl K, McElroy T, Katahoire A. Effectiveness of a SEL/Mindfulness Program on Northern Ugandan Children. Int J Sch Educ Psychol. 2020;

62. Ambelu A, Mulu T, Seyoum A, Ayalew L, Hildrew S. Resilience Dynamics after Interventions Made Among School Children of Rural Ethiopia. Heliyon. 2019;5(e01464).

63. Tudor K, Maloney S, Raja A, Baer R, Blakemore SJ, Byford S, et al. Universal Mindfulness Training in Schools for Adolescents: a Scoping Review and Conceptual Model of Moderators, Mediators, and Implementation Factors. Prev Sci [Internet]. 2022;23(6):934–53. Available from: https://doi.org/10.1007/s11121-022-01361-9

64. Draper-Clarke LJ. Compassion-based Mindfulness Training in Teacher Education: The Impact on Student Teachers at a South African University. South African J High Educ. 2020;34(1):57–79.

65. Galvin M, Byansi W. A Systematic Review of Task Shifting for Mental Health in sub-Saharan Africa. Int J Ment Health. 2020 Oct;49(4):336–60.

66. Developing and Delivering A Mindfulness Intervention [Internet]. Centre for Global Development. 2023 [cited 2023 May 8]. Available from: https://www.abdn.ac.uk/education/research/cgd/nihr-camw-subsaharan-africa/developing-delivering-a-mindfulness-intervention-1828.php#panel1833

67. Musanje K, Camlin CS, Kamya MR, Vanderplasschen W, Louise Sinclair D, Getahun M, et al. Culturally adapting a mindfulness and acceptance-based intervention to support the mental health of adolescents on antiretroviral therapy in Uganda. PLOS Glob Public Heal. 2023;3(3):e0001605.

68. Shapiro S, Rechtschaffen D, Sousa S de. Handbook of Mindfulness in Education. In: Schonert-Reichl KA, Roeser RW, editors. Handbook of Mindfulness in Education Integrating Theory and Research into Practice [Internet]. Verlag New York: Springer; 2016. Available from: https://link.springer.com/book/10.1007/978-1-4939-3506-i

69. Dunning D, Tudor K, Radley L, Dalrymple N, Funk J, Vainre M, et al. Do mindfulness-based programmes improve the cognitive skills, behaviour and mental health of children and adolescents? An updated meta--analysis of randomised controlled trials. BMJ Evid Based Ment Heal [Internet]. 2022;25(3):135–42. Available from: https://ebmh.bmj.com/content/25/3/135

70. Kuyken W, Ball S, Crane C, Ganguli P, Jones B, Montero-Marin J, et al. Effectiveness and cost-effectiveness of universal school-based mindfulness training compared with normal school provision in reducing risk of mental health problems and promoting wellbeing in adolescence: The MYRIAD cluster randomised controlled trial. Evid Based Ment Health [Internet]. 2022;25(3):99–109. Available from: https://ebmh.bmj.com/content/ebmental/25/3/99.full.pdf

71. Kuyken W, Ball S, Crane C, Ganguli P, Jones B, Montero-Marin J, et al. Effectiveness of universal school-based mindfulness training compared with normal school provision on teacher mental health and school climate: Results of the MYRIAD cluster randomised controlled trial. Evid Based Ment Health. 2022;25(3):125–34.

72. Thabane L, Lancaster G. A guide to the reporting of protocols of pilot and feasibility trials. Pilot Feasibility Stud. 2019;5(1):5–7.

73. Bhaskar R, Danermark B, Price L. Interdisciplinarity and Wellbeing: A Critical Realist General Theory of Interdisciplinarity [Internet]. London and New York, NY: Routledge; 2018. Available from: https://www.amazon.co.uk/Interdisciplinarity-Wellbeing-Critical-Realist-Routledge/dp/0415403715/ref=sr_1_1?crid=4O3D8WANEIEM&keywords=critical+realism+wellbeing&qid=1662303252&s=books&sprefix=critical+realism+wellbeing+%2Cstripbooks%2C70&sr=1-1

74. Bhaskar R. The Possibility of Naturalism. A Philosophical Critique of the Contemporary Human Sciences. Hassocks: Harvester Press; 1989.

75. Archer M. Realist Social Theory: The Morphogenetic Approach. Cambridge: Cambridge University Press; 2008.

76. Bhaskar R. Enlightened Common Sense: The Philosophy of Critical Realism [Internet]. Abingdon: Routledge; 2016. Available from: https://www.amazon.co.uk/Enlightened-Common-Sense-Ontological-Explorations/dp/0415583799/ref=sr_1_1?keywords=Enlightened+Common+Sense%3A+The+Philosophy+of+Critical+Realism&qid=1646410466&sr=8-1

77. Imel Z, Baldwin S, Bonus K, Maccoon D. Beyond the individual: group effects in mindfulness-based stress reduction. Psychother Res [Internet]. 2008;18(6):735–42. Available from: https://pubmed.ncbi.nlm.nih.gov/18815948/

78. Skinner E, Beers J. Mindfulness and Teachers’ Coping in the Classroom: A Developmental Model of Teacher Stress, Coping, and Everyday Resilience. In: Schonert-Reichl KA, Roeser RW, editors. Handbook of Mindfulness in EducationIntegrating Theory and Research into Practice [Internet]. Verlag New York: Springer; 2016. Available from: https://link.springer.com/book/10.1007/978-1-4939-3506-2?code=8eb2f0dd-ebed-4d00-9bc8-ab98d5cb29a6

79. Larson KE, Nguyen AJ, Orozco Solis MG, Humphreys A, Bradshaw CP, Lindstrom Johnson S. A systematic literature review of school climate in low and middle income countries. Int J Educ Res. 2020;102(June).

80. Tarrasch R. Mindful Schooling: Better Attention Regulation Among Elementary School Children Who Practice Mindfulness as Part of their School Policy. J Cogn Enhanc. 2017;1(2):84–95.

81. Zenner C, Herrnleben-Kurz S, Walach H. Mindfulness-based interventions in schools-a systematic review and meta-analysis. Front Psychol [Internet]. 2014;5(603):1–20. Available from: https://www.ncbi.nlm.nih.gov/pmc/articles/PMC4075476/pdf/fpsyg-05-00603.pdf

82. Ritchhart R, Perkins DNN. Life in the mindful classroom: Nurturing the disposition of mindfulness. J Soc Issues. 2000;56(1):27–47.

83. Bonell C, Fletcher A, Jamal F, Wells H, Harden A, Murphy S, et al. Theories of how the school environment impacts on student health: Systematic review and synthesis. Heal Place [Internet]. 2013;24:242–9. Available from: http://dx.doi.org/10.1016/j.healthplace.2013.09.014

84. Wang M Te, Degol JL. School Climate: a Review of the Construct, Measurement, and Impact on Student Outcomes [Internet]. Vol. 28, Educational Psychology Review. Educational Psychology Review; 2016. 315–352 p. Available from: http://dx.doi.org/10.1007/s10648-015-9319-1

85. Gray C, Wilcox G, Nordstokke D. Teacher Mental Health, School Climate, Inclusive Education and Student Learning: A Review. Vol. 58, Canadian Psychology. 2017. p. 203–10.

86. Thapa A, Cohen J, Guffey S, Higgins-D’Alessandro A. A Review of Research. Rev Educ Res. 2013;83(3):357–85.

87. Jamal F, Fletcher A, Harden A, Wells H, Thomas J, Bonell C. The school environment and student health: A systematic review and meta-ethnography of qualitative research. BMC Public Health. 2013;13(1):1–11.

88. Bonell C, Mathiot A, Allen E, Bevilacqua L, Christie D, Elbourne D, et al. Initiating change locally in bullying and aggression through the school environment (INCLUSIVE) trial: Update to cluster randomised controlled trial protocol. Trials. 2017;18(1):1–14.

89. Abbott P, Stanley I, Nixson G, D’Ambruoso L. A protocol for asystematic critical realist synthesis of school mindfulness interventions designed to promote pupils’ mental wellbeing. CRD42023410484 [Internet]. PROSPERO. 2023 [cited 2023 Apr 5]. Available from: https://www.crd.york.ac.uk/PROSPERO/#searchadvanced

90. Abbott P, Stanley I, Nixson G, D’Ambruoso L. A protocol for asystematic realist synthesis of school mindfulness interventions designed to promote mental wellbeing. medRxiv [Internet]. 2023;MEDRXIV/20. Available from: https://www.medrxiv.org/content/10.1101/2023.04.06.23288255v1

91. Abbott P, Shanks R, Stanley I, D’Ambruoso L. A protocol for a critical realist systematic synthesis of school climate change interventions designed to promote pupils’ wellbeing in Low-and Middle-Income Countries. CRD42023417735 [Internet]. PROSPERO. 2023 [cited 2023 May 8]. Available from: https://www.crd.york.ac.uk/prospero/#myprospero

92. Participatory Action Research [Internet]. Centre for Global Development. 2023 [cited 2023 May 8]. Available from: https://www.abdn.ac.uk/education/research/cgd/nihr-camw-subsaharan-africa/open-access-training-1735.php#panel1848

93. Rapid Research Evaluation and Appraisal Lab [Internet]. [cited 2023 May 9]. Available from: https://www.rapidresearchandevaluation.com/

94. Bonell C, Fletcher A, Morton M, Lorenc T, Moore L. Methods don’t make assumptions, researchers do: A response to Marchal etal. Soc Sci Med [Internet]. 2013;94:81–2. Available from: http://dx.doi.org/10.1016/j.socscimed.2013.06.026

95. Hammersley M. The relationship between qualitative and quantitative research: paradigm loyalty versus eclecticism. In: Richardson JTE, editor. Handbook of Qualitative Research Methods for Psychology and the Social Sciences. Leicester: British Psychological Society; 1996.

96. Maxwell JA, Mittapalli K. Realism as a stance for mixed methods research. In: Tashakkori A, Teddlie C, editors. Sage Handbook of Mixed Methods in Social and Behavioural Research. 2nd ed. London: Sage; 2010.

97. Mingers J. A critique of statistical modelling in management science from a critical realist perspective: Its role within multimethodology. J Oper Res Soc. 2006;57(2):202–19.

98. Mcevoy P, Richards D. A critical realist rationale for using a combination of quantitative and qualitative methods. J Res Nurs. 2006;11(1):66–78.

99. Modell S. In defence of triangulation: A critical realist approach to mixed methods research in management accounting. Manag Account Res. 2009;20(3):208–21.

100. Mukumbang F. Retroductive Theorizing: A Contribution of Critical Realism to Mixed Methods Research. J Mix Methods Res [Internet]. 2023;17(1):93–114. Available from: https://doi.org/10.1177/15586898211049847

101. Brönnimann A. How to phrase critical realist interview questions in applied social science research. J Crit Realis. 2022;21(1):1–24.

102. Sayer A. Method in Social Science: A Realist Approach [Internet]. 2nd ed. London: Routledge; 2010. Available from: https://www.amazon.co.uk/Method-Social-Science-Revised-2nd/dp/0415582474/ref=sr_1_8?crid=36VCTDY04BZQP&keywords=andrew+sayer&qid=1646414830&sprefix=Andrew+Sayer%2Caps%2C58&sr=8-8

103. Cantril’s Ladder [Internet]. [cited 2023 Apr 9]. Available from: https://www.ncbi.nlm.nih.gov/books/NBK189562/

104. Beyond Blue School Climate Survey [Internet]. Available from: https://www.google.co.uk/search?q=beyond+blue+school+climate+questionnaire+pupil+version&ei=UcgyZMKRA5PngAbCubnwBA&ved=0ahUKEwiC_tqI_pz-AhWTM8AKHcJcDk4Q4dUDCA8&uact=5&oq=beyond+blue+school+climate+questionnaire+pupil+version&gs_lcp=Cgxnd3Mtd2l6LXNlcnAQAzI

105. In School Behaviour Survey [Internet]. [cited 2023 Apr 9]. Available from: https://tombennetttraining.co.uk/wp-content/uploads/2021/04/The-Bennett-Behaviour-Survey.pdf

106. Child and Adolescent Mindfulness Measure [Internet]. Available from: https://www.ruthbaer.com/academics/CAMM.pdf

107. Kessler Psychological Distress Scale (K10) [Internet]. [cited 2023 Apr 9]. Available from: https://www.tac.vic.gov.au/files-to-move/media/upload/k10_english.pdf

108. Teacher Job Satisfaction Questionnaire [Internet]. [cited 2023 Apr 9]. Available from: https://www.google.co.uk/search?q=questionnaire+teacher+job+satisfaction.doc&ei=btEyZIyBO4zvgQaUnaDoCQ&oq=Questionnaire+Teacher+satisfaction+&gs_lcp=Cgxnd3Mtd2l6LXNlcnAQARgAMgYIABAWEB4yBggAEBYQHjIICAAQigUQhgMyCAgAEIoFEIYDMggIABCKBRCGAzIICAAQigUQhgNKBAhBGA

109. Child Behaviour Questionnaire (parents’ version) [Internet]. [cited 2023 Apr 9]. Available from: https://www.bristol.ac.uk/media-library/sites/social-community-medicine/migrated/documents/probit2sdqparent.pdf

110. Wiltshire G, Ronkainen N. A realist approach to thematic analysis: making sense of qualitative data through experiential, inferential and dispositional themes. J Crit Realis [Internet]. 2021;20(2):159–80. Available from: https://doi.org/10.1080/14767430.2021.1894909

111. Hatt K. View of Considering Complexity_ Toward A Strategy for Non-linear Analysis. Can J Sociol. 2009;34(2):313–47.

112. Hinds K, Dickson K. Realist synthesis: a critique and an alternative. J Crit Realis [Internet]. 2021;20(1):1–17. Available from: https://doi.org/10.1080/14767430.2020.1860425

113. Bellazzecca E, Teasdale S, Biosca O, Skelton DA. The health impacts of place-based creative programmes on older adults’ health: A critical realist review. Heal Place [Internet]. 2022;76(January):102839. Available from: https://doi.org/10.1016/j.healthplace.2022.102839

114. Mukumbang F, De Souza D, Liu H, Uribe G, Moore C, Fotheringham P, et al. Unpacking the design, implementation and uptake of community-integrated health care services: A critical realist synthesis. BMJ Glob Heal [Internet]. 2022;7(8):1–14. Available from: https://gh.bmj.com/content/bmjgh/7/8/e009129.full.pdf

115. Melendez-Torres GJ, Warren E, Viner R, Allen E, Bonell C. Moderated mediation analyses to assess intervention mechanisms for impacts on victimisation, psychosocial problems and mental wellbeing: Evidence from the INCLUSIVE realist randomised trial. Soc Sci Med [Internet]. 2021;279(May):113984. Available from: https://doi.org/10.1016/j.socscimed.2021.113984

116. Pituch KA, Stapleton LM. Distinguishing Between Cross-and Cluster-Level Mediation Processes in the Cluster Randomized Trial. Sociol Methods Res [Internet]. 2012;41(4):630–70. Available from: https://journals.sagepub.com/doi/epub/10.1177/0049124112460380

117. Hoddy ET. Critical realism in empirical research: employing techniques from grounded theory methodology. Int J Soc Res Methodol. 2019;22(1):111–24.

118. Oliver C. Critical realist grounded theory: A new approach for social work research. Br J Soc Work. 2012;42(2):371–87.

119. Brooks J, McCluskey S, Turley E, King N. The Utility of Template Analysis in Qualitative Psychology Research. Qual Res Psychol. 2015;12(2):202–22.

120. NIHR Global Health Research Group on promoting children’s and adolescents’ mental wellbeing in sub-Saharan Africa. About the Project [Internet]. Policies on Safeguarding and Whistleblowing. 2023. Available from: https://www.abdn.ac.uk/education/research/cgd/nihr-camw-subsaharan-africa/about-the-project-1616.php

121. Centre for Global Development. Open Access Training [Internet]. 2013 [cited 2023 Apr 7]. Available from: https://www.abdn.ac.uk/education/research/cgd/nihr-camw-subsaharanafrica/open-access-training-1735.php

122. Publications Policy [Internet]. Centre for Global Development. 2023 [cited 2023 May 8]. Available from: https://www.abdn.ac.uk/education/documents/PublicationsPolicy_2023.04.17_withCS.pdf

