## Supplementary Table 1 for "Study protocol for a Critical Realist pilot cluster-randomised controlled trial of a whole-school-based mindfulness intervention (SBMI) promoting child and adolescent mental wellbeing in Rwanda and Ethiopia"

Additional Material 1

Sources of Evidence and Examples of Research Questions in Phase 2

| **Research Questions** | **Participants** | **Survey Questionnaire** | **Psychometric tools** | **Qualitative interviews and FGDs** | **Routine Administrative Data** | **0bservation by researchers** | **Cost Analysis** |
|---|---|---|---|---|---|---|---|
| **What is the school and community climate like?** | Baseline **(yr. 2/yr3)** | | | | | | |
| | CAs | | School climate<br>Life satisfaction<br>Mental wellbeing<br>Mindfulness<br>Emotional regulation<br>Resilience<br>School engagement<br>Pupil behaviour<br>School satisfaction | Please tell me about what your school is like.<br>How do you get on with your classmates?<br>How do you get on with your teacher?<br>What do you like and what do you dislike about your school?<br>How do CAs behave in the classroom?<br>How do teachers maintain discipline in the classroom?<br>Tell me about how your friends influence how your behave? | | How children behaviour in class and on the playground<br>How teachers interact with CAs in school<br>How teachers interact with children in the classroom.<br>How children behave in class<br>How teachers maintain discipline | |
| | Teachers | | Mental wellbeing | Please tell me what it is like to be a | | | |

| Research Questions | Participants | Survey Questionnaire | Psychometric tools | Qualitative interviews and FGDs | Routine Administrative Data | 0bservation by researchers | Cost Analysis |
|---|---|---|---|---|---|---|---|
| | | | Job satisfaction<br>Life satisfaction<br>School climate<br>Pupil behaviour | teacher at this school.<br>Tell me about how CAs behave in class.<br>What sort of CAs have behavioural/attention problems?<br>What sort of CAs need psychological support?<br>Tell me about CAs attitude to school<br>How has the school organisation influenced how you have taught?<br>Why has it had that Influence.<br>How does the school organisation influence the behaviour of pupils?<br>Why does it have the influence? | | | |

| Research Questions | Participants | Survey Questionnaire | Psychometric tools | Qualitative interviews and FGDs | Routine Administrative Data | 0bservation by researchers | Cost Analysis |
|---|---|---|---|---|---|---|---|
| | | | | How do you maintain discipline in your classroom?<br>Why do you use these methods to maintain discipline? | | | |
| | School Administration | | School climate<br>Pupil behaviour | | CA attendance and dropout rates, examination results.<br>Teacher absence. | | |
| | Mothers/primary caregivers | Lived Poverty Scale (AfroBarometer)<br>Wealth Index (DHS)<br>Child relationship with parents, siblings, peer group, attitude to school, relationship with teachers, relationship with other pupils. | Mental wellbeing<br>Satisfaction with Family life<br>Life satisfaction | Please tell me about the primary school your child/children attend.<br><br>Tell me about how the teachers treat the children.<br><br>Tell me about your children's attitude to school | | | |

| Research Questions | Participants | Survey Questionnaire | Psychometric tools | Qualitative interviews and FGDs | Routine Administrative Data | 0bservation by researchers | Cost Analysis |
|---|---|---|---|---|---|---|---|
| | | Relationships with members of the community.<br>Behaviour at home<br>How parents discipline their children | | Tell me about your children's attitudes to their teachers<br><br>Tell me about the relationship between the school and parents.<br><br>Tell me about how your child behaves at home?<br>Please tell me about how you discipline your child at home? | | | |
| | **Process evaluation (yr. 3)** | | | | | | |
| | CA | | School climate<br>Life satisfaction<br>Mental wellbeing<br>Mindfulness<br>Emotional regulation | Please tell me about what has happened at school since the start of the school year.<br>Why do you think that these changes have taken place? | | How children behaviour in class and on the playground<br>How teachers interact with | |

| Research Questions | Participants | Survey Questionnaire | Psychometric tools | Qualitative interviews and FGDs | Routine Administrative Data | 0bservation by researchers | Cost Analysis |
|---|---|---|---|---|---|---|---|
| | | | Resilience<br>School engagement<br>Pupil behaviour<br>School satisfaction | Please tell me about any changes in children's behaviour at your school since the beginning of the school year.<br>How has their behaviour changed?<br>How has your behaviour changed?<br>Why do you think that there have been these changes<br>Tell me about any changes in how teachers treat the children in your school<br>Tell me about any changes in the way teachers manage discipline in the classroom<br>Tell me why you think that these changes have taken place. | | CAs in school<br>How teachers interact with children in the classroom.<br>How children behave in class<br>How teachers maintain discipline<br><br>How CAs respond to mindfulness training | |

| Research Questions | Participants | Survey Questionnaire | Psychometric tools | Qualitative interviews and FGDs | Routine Administrative Data | 0bservation by researchers | Cost Analysis |
|---|---|---|---|---|---|---|---|
| | | | | Tell me about the mindfulness exercises you have been taught.<br>Tell me about practising the mindfulness exercises<br>Tell me about things that have made it difficult for you to do the mindfulness exercises.<br>Can you think of anything else other than the mindfulness exercises that could have brought about the changes you have observed? | | | |
| | Teachers | | Mental wellbeing<br>Job satisfaction<br>Life satisfaction<br>School climate | Please tell me about any changes in the behaviour of pupils in your class since the start of the school year | | | |

| Research Questions | Participants | Survey Questionnaire | Psychometric tools | Qualitative interviews and FGDs | Routine Administrative Data | 0bservation by researchers | Cost Analysis |
|---|---|---|---|---|---|---|---|
| | | | Pupil behaviour | How have they changed<br>Why do you think that these changes have taken place?<br>How has your behaviour changed since the beginning of the school year?<br>Why do you think that these changes have taken place?<br>When you reflect on the mindfulness intervention, what motivated you to implement it?<br>How has the mindfulness intervention changed the pupils in your class?<br>Why do you think that it has brought about these changes? | | | |

| Research Questions | Participants | Survey Questionnaire | Psychometric tools | Qualitative interviews and FGDs | Routine Administrative Data | 0bservation by researchers | Cost Analysis |
|---|---|---|---|---|---|---|---|
| | | | | Can you think of anything else, other than the mindfulness exercises that could have brought about these changes? | | | |
| | School Administration | | School climate<br>Pupil behaviour | | CA attendance and dropout rates, examination results.<br>Teacher absence. | | |
| | Mothers/primary caregivers | | | | | | |
| | **End-of-line (yr. 3** | | | | | | |
| | CAs | | School climate<br>Life satisfaction<br>Mental wellbeing<br>Mindfulness<br>Emotional regulation<br>Resilience | Please tell me about what has happened at school since the start of the school year.<br>Why do you think that these changes have taken place? | | How children behaviour in class and on the playground<br>How teachers interact with | |

| **Research Questions** | **Participants** | **Survey Questionnaire** | **Psychometric tools** | **Qualitative interviews and FGDs** | **Routine Administrative Data** | **0bservation by researchers** | **Cost Analysis** |
|---|---|---|---|---|---|---|---|
| | | | School engagement<br>Pupil behaviour<br>School satisfaction | Please tell me about any changes in children's behaviour at your school since the beginning of the school year.<br>How has their behaviour changed?<br>How has your behaviour changed?<br>Why do you think that there have been these changes<br>Tell me about any changes in how teachers treat the children in your school<br>Tell me why you think that these changes have taken place.<br>Tell me about the mindfulness exercises you have been taught. | | CAs in school<br>How teachers interact with children in the classroom.<br>How children behave in class<br>How teachers maintain discipline<br><br>How teachers teach the mindfulness exercises<br><br>How CAs respond to mindfulness training | |

| **Research Questions** | **Participants** | **Survey Questionnaire** | **Psychometric tools** | **Qualitative interviews and FGDs** | **Routine Administrative Data** | **0bservation by researchers** | **Cost Analysis** |
|---|---|---|---|---|---|---|---|
| | | | | Tell me about practising the mindfulness exercises | | | |
| | Teachers | | Mental wellbeing<br>Job satisfaction<br>Life satisfaction<br>School climate<br>Pupil behaviour | Please tell me about any changes in the behaviour of pupils in your class since the start of the school year<br>How have they changed<br>Why do you think that these changes have taken place?<br>How has your behaviour changed since the beginning of the school year?<br>Why do you think that these changes have taken place?<br>When you reflect on the mindfulness intervention, what motivated you to implement it? | | | |

| **Research Questions** | **Participants** | **Survey Questionnaire** | **Psychometric tools** | **Qualitative interviews and FGDs** | **Routine Administrative Data** | **0bservation by researchers** | **Cost Analysis** |
|---|---|---|---|---|---|---|---|
| | | | | How has the mindfulness intervention changed the pupils in your class?<br>Why do you think that it has brought about these changes? | | | |
| | School Administration | | School climate<br>Pupil behaviour | | CA attendance and dropout rates, examination results.<br>Teacher absence. | | |
| | Mothers/primary carergivers | Lived Poverty Scale (AfroBarometer)<br>Wealth Index (DHS)<br>Child anxious, depressed, has difficulty in sleeping, aggressive, relationships with | Mental wellbeing<br>Satisfaction with Family life<br>Life satisfaction | What changes have you noticed in your child/children over the last school year<br>Why do you think that these changes have occurred? | | | |

| Research Questions | Participants | Survey Questionnaire | Psychometric tools | Qualitative interviews and FGDs | Routine Administrative Data | 0bservation by researchers | Cost Analysis |
|---|---|---|---|---|---|---|---|
| | | parents, siblings, peer groups, attitude to school, relationship with teachers and with other pupils Parents' attitude to school. Compared with the beginning of the school year, would you say that your child's behaviour at home is a lot better, better, much the same, worse, or a lot worse? | | | | | |
| **What is the status of the mental** | **Baseline (yr. 2)** | | | | | | |
| | CAs | | Mental wellbeing Mindfulness | | How do you feel about school and | | |

| **Research Questions** | **Participants** | **Survey Questionnaire** | **Psychometric tools** | **Qualitative interviews and FGDs** | **Routine Administrative Data** | **0bservation by researchers** | **Cost Analysis** |
|---|---|---|---|---|---|---|---|
| **wellbeing of CAs?** | | | Emotional regulation<br>Resilience | | your life in general?<br>Why do you think that you feel like that? | | |
| | Teachers | | | | What do you think are the main health problems CAs at your Schools have?<br>What is the mental wellbeing like of the CAs in your class?<br>Why do you think the mental wellbeing of children in your class is as it is? | | |
| | School Administration | | | | | | |
| | Mothers/primary caregivers | Child anxious, depressed, has difficulty in | | | What do you think are the main health | | |

| **Research Questions** | **Participants** | **Survey Questionnaire** | **Psychometric tools** | **Qualitative interviews and FGDs** | **Routine Administrative Data** | **0bservation by researchers** | **Cost Analysis** |
|---|---|---|---|---|---|---|---|
| | | sleeping, aggressive. | | | problems that CAs have in this community? What do you think about the mental wellbeing of CAs? Why do you think this? Why do some CAs have poor mental wellbeing? | | |
| | Policy actors | | | What are the main health problems for CAs in Rwanda? Why are these a problem? How much of a problem is mental wellbeing? Why do you think that some CAs have poor mental wellbeing? | | | |

| Research Questions | Participants | Survey Questionnaire | Psychometric tools | Qualitative interviews and FGDs | Routine Administrative Data | 0bservation by researchers | Cost Analysis |
|---|---|---|---|---|---|---|---|
| | | | | Why do you think it is a problem? | | | |
| | **Process evaluation (yr. 3)** | | | | | | |
| | CAs | | Mental wellbeing<br>Mindfulness<br>Emotional regulation<br>Resilience | | Have you noticed any changes in how you feel about school and your life since the beginning of the school year?<br>Why do you think that these changes have happened? | | |
| | Teachers | | | | | | |
| | School Administration | | | | | | |
| | Mothers/primary caregivers | | | | | | |
| | Policy actors | | | | | | |

| Research Questions | Participants | Survey Questionnaire | Psychometric tools | Qualitative interviews and FGDs | Routine Administrative Data | 0bservation by researchers | Cost Analysis |
|---|---|---|---|---|---|---|---|
| | **End-of-line (yr. 3** | | | | | | |
| | CAs | | Mental wellbeing<br>Mindfulness<br>Emotional regulation<br>Resilience | | Have you noticed any changes in how your fellow pupils feel about school and their life in general since the beginning of the school year?<br><br>Have you noticed any changes in how you feel about school and your life since the beginning of the school year?<br>Why do you think that these changes have happened? | | |

| Research Questions | Participants | Survey Questionnaire | Psychometric tools | Qualitative interviews and FGDs | Routine Administrative Data | 0bservation by researchers | Cost Analysis |
|---|---|---|---|---|---|---|---|
| | Teachers | | | | | | |
| | School Administration | | | | | | |
| | Mothers/primary caregivers | Child anxious, depressed, has difficulty in sleeping, aggressive. | | | | | |
| **For whom, how and why were there these changes in CA's mental wellbeing after eight months of delivery** | **Baseline (yr. 2)** | | | | | | |
| | CAs | Demographic data - age, gender, class at school, location (rural Rwanda, urban Ethiopia). Have you been practising mindfulness exercises? | | | | | |
| | Teachers | | | | | | |
| | School Administration | | | | | | |
| | Mothers/primary caregivers | Demographic data - age of CAs attending primary school, | | | | | |

| Research Questions | Participants | Survey Questionnaire | Psychometric tools | Qualitative interviews and FGDs | Routine Administrative Data | 0bservation by researchers | Cost Analysis |
|---|---|---|---|---|---|---|---|
| | | socioeconomic status of household (Wealth Index, Lived Poverty Scale) | | | | | |
| | **Process evaluation (yr. 3)** | | | | | | |
| | CAs | | | | | | |
| | Teachers | | | | | | |
| | School Administration | | | | | | |
| | Mothers/primary caregivers | | | | | | |
| | **End-of-line (yr. 3** | | | | | | |
| | CAs | | | | | | |
| | Teachers | | | | | | |
| | School Administration | | | | | | |
| | Mothers/primary caregivers | | | | | | |
| | Policy actors | | | | | | |

| Research Questions | Participants | Survey Questionnaire | Psychometric tools | Qualitative interviews and FGDs | Routine Administrative Data | 0bservation by researchers | Cost Analysis |
|---|---|---|---|---|---|---|---|
| **Is the intervention culturally acceptable, affordable, and cost-effective** | **Baseline (yr. 2)** | | | | | | |
| | CAs | | | | | | |
| | Teachers | | | | What is your attitude to teaching mindfulness to your class? Why do you say that? | | |
| | School Administration | | | | | | |
| | Mothers/primary caregivers | | | | | | |
| | Policy actors | | | | What is your attitude to CAs being taught mindfulness exercises in school? Why do you say that? | | |
| | **Process evaluation (yr. 3)** | | | | | | |
| | CAs | | | | What do you think of the | | |

| Research Questions | Participants | Survey Questionnaire | Psychometric tools | Qualitative interviews and FGDs | Routine Administrative Data | 0bservation by researchers | Cost Analysis |
|---|---|---|---|---|---|---|---|
| | | | | | mindfulness exercises you have been taught at school? Why do you feel this way about them? | | |
| | Teachers | | | | What do you think about teaching mindfulness exercises? Why do you feel this way about teaching them to the children in your class? | | |
| | School Administration | | | | | | |
| | Mothers/primary carregivers | | | | | | |
| | **End-of-line (yr. 3** | | | | | | |

| **Research Questions** | **Participants** | **Survey Questionnaire** | **Psychometric tools** | **Qualitative interviews and FGDs** | **Routine Administrative Data** | **0bservation by researchers** | **Cost Analysis** |
|---|---|---|---|---|---|---|---|
| | CAs | Where have you been practising the mindfulness exercises? | | | What do you think of the mindfulness exercises you have been taught at school?<br>Why do you feel this way about them?<br>How have you used the mindfulness exercises? | | |
| | Teachers | | | | What do you think about teaching mindfulness exercises?<br>Why do you feel this way about teaching them to the children in your class? | | |
| | School Administration | | | | | | |

| Research Questions | Participants | Survey Questionnaire | Psychometric tools | Qualitative interviews and FGDs | Routine Administrative Data | 0bservation by researchers | Cost Analysis |
|---|---|---|---|---|---|---|---|
| | Mothers/primary caregivers | Have your child/children told you about the mindfulness exercises they have been taught at school this year?<br><br>Do your children practice mindfulness exercises at home?<br><br>What is your attitude to your children being taught mindfulness?<br><br>Why do you have these attitudes? | | | What is your attitude to the exercises your children have been taught in school?<br>Why do you have this attitude towards them?<br>What do your children say about the mindfulness exercises they are taught at school? | | |
| | Policy actors | | | | What is your attitude to CAs being taught mindfulness | | |

| Research Questions | Participants | Survey Questionnaire | Psychometric tools | Qualitative interviews and FGDs | Routine Administrative Data | 0bservation by researchers | Cost Analysis |
|---|---|---|---|---|---|---|---|
| | | | | | exercises in school? Why do you have this attitude? Can you think of any alternative explanations for the changes that have occurred in the intervention schools? | | |
